# Global risk assessment of Lyme borreliosis transmission

**DOI:** 10.1101/2025.01.29.25321312

**Authors:** Marina Cobos-Mayo, Adrián Martín-Taboada, Alisa Aliaga-Samanez, Marina Segura, Jesús Olivero

## Abstract

We analysed the geographic risk of Lyme borreliosis taking into account the biogeography of tick vectors and carrier hosts, together with environmental and anthropogenic factors. Four pathogeographical scenarios were set in order to represent the contribution of vectors and hosts in the spatial zoonotic risk. For that propose, we built distribution models based on the occurrence of Lyme borreliosis cases in humans and ixodid vectors. Besides *Ixodes* species, we considered other ixodid ticks with potential to be vectors. These models were combined through fuzzy logic operators, according to the criteria stablished in each scenario. Finally, the transmission risk model for Lyme borreliosis which best fitted its global distribution was selected. The risk model selected considered ixodid vectors and mammal carriers as explanatory variables together with environment and anthropogenic factors. *Ixodes* species contributed to explain the geographical risk of Lyme borreliosis to larger extent than other ixodid ticks. The risk model described regions with Lyme borreliosis transmission risk where its presence is still uncertain, such as northern Africa and inland areas of western USA. Likewise, our model indicated favourable conditions for the presence of human cases in northern latitudes beyond its endemic distribution. Applying this multi-scenario methodology approach have led us to a risk model, in which the diversity of ixodid vectors and carrier hosts might modify the spatial risk without a geographical limitation.

## 1. INTRODUCTION

Lyme borreliosis (LB) is caused by several bacteria species from the *Borrelia burgdorferi* sensu lato (s.l.) complex, and is transmitted through the bite of ixodid ticks, mainly of *Ixodes* species (Stanek et al., 2012). Although LB is the most prevalent tick-borne disease in the Holarctic, the overall epidemiological situation is rather uncertain due to the lack of standardised diagnosis procedures and of a robust surveillance system (Blanchard et al., 2022). So far, only one vaccine is currently under development (Burn et al., 2023).

Few works have assessed the geographical risk of LB at a large scale, and no one has considered both vectors and reservoirs involved as predictors explaining spatial risk (Chumachenko et al., 2022; Ozdenerol et al., 2015). This situation is mainly due to a lack of high-resolution epidemiological data and to the complexity of its zoonotic cycle (Braks et al., 2016), which involves multiple interactions between tick vectors and a wide range of carrier hosts, i.e., reservoir and non-reservoir hosts (Kahl et al., 2002). The relevant wild carriers of *B. burgdorferi* s.l. have been confirmed to be small mammals, mostly rodent species (Steinbrink et al., 2022). Some species from Lagomorpha and Eulipotyphla orders, such as hedgehogs and squirrels, are also identified as wild carriers in enzootic cycles (Kurtenbach et al., 2002; Piesman & Gern, 2004). Moreover, although medium to large-size mammals such as deer are not carriers of *Borrelia* species, they are main tick vector hosts. Nymph and adult ticks get infected with *B. burgdorferi* s.l., through co-feeding transmission while they are feeding on deer (Voordouw, 2015). Thus, the presence of deer favours the maintenance of tick populations and the connection between them, and so the circulation of the pathogen in endemic areas (van Wieren & Hofmeester, 2016). For that reason, deer are also considered amplifier hosts in the LB zoonotic cycle (Kahl et al., 2002).

Considering a pathogeographical approach in the analysis of geographic disease risk allows us to relate the distribution of reported human cases to the biogeography of vector and carrier species, and also to the presence of biotic and abiotic conditions that favour the transmission of pathogens to humans (Murray et al., 2018). This methodological framework has been successfully applied to assess the spatio-temporal dynamics of mosquito-borne diseases (Aliaga-Samanez et al., 2021, 2022), which provides a robust basis for analysing the spatial risk of tick-borne disease at a global level. In Aliaga-Samanez *et al*.’s (2021, 2022) models, the presence of vectors was assumed to be needed, and so to act as a geographical limiting factor for pathogen transmission; whereas the presence of non-human animal carriers was assumed to increase, but not limit transmission risk in some regions of the World. In the case of LB, vectors and wild carriers are certain agents for disease transmission in the ecosystems, but could be considered to be virtually ubiquitous in geographical terms. This being the case, ticks and carriers but could not be acting as geographically limiting factors. The way how vectors and carriers condition the spatial zoonotic risk could give place to different pathogeographic scenarios, according to which zoonotic agents behave or not as a geographically limiting factors. In order to check on this regarding the LB transmission risk, in this work we have produced alternative models based on different hypothetical pathogeographic scenarios. Finally, we focused on the scenario that produced the best fitted model, and used it to build the highest-resolution global map of LB transmission risk in humans up to now.

## 2. MATERIALS AND METHODS

### 2.1 General methodological framework

We established four different pathogeographic scenarios, each one leading to a favourability model that described favourable areas for the transmission of LB to occur (i.e., a “transmission risk model”, TM): 1) both vectors and wild carriers behave as geographically limiting factors in shaping the area at risk of zoonotic transmission (VCL scenario); 2) only wild carriers are limiting factors (CL scenario); 3) only vectors are limiting factors (VL scenario); and 4) neither vectors nor wild carriers are limiting factors, i.e. vector and wild carriers are mutually compensating factors together with other environmental factors (NL scenario). The existence of a geographically limiting factor involved the spatial intersection between the distributions of the agents limiting the transmission risk, so that favourable conditions for every limiting agent to occur in a location be warrantied.

The modelling approach considered here was based on the Favourability Function (Acevedo & Real, 2012; Real et al., 2006) (see section 2.3), whose output represents a measure of which level environmental conditions at a given location are favourable to the occurrence of the modelled entity. Such a level is equivalent to the degree of membership in a fuzzy set of favourable locations (Barbosa & Real, 2012), and so fuzzy logic operators can be used for combining favourability models. In this context, on the one hand, a model considering limiting zoonotic agents (such as vectors or carriers) can be built as the combination of different models, each one based on a different agent, through a fuzzy intersection (Romero et al., 2016). The fuzzy intersection consists of retaining, for every location, the lowest degree of membership (i.e., the minimum favourability value) recorded by the models to be combined (Zadeh, 1965). On the other hand, zoonotic agents not acting as geographically limiting factors for disease transmission could still be considered as predictor variables within a model. This would represent a chance for these agents, together with other environmental variables, to increase or decrease the level of transmission risk in the area. The predictive relevance of these non-limiting agents was weighted through adjustments of their coefficients in the model equation.

We proceeded by building “vector models” (VM) using, as dependent variables, the occurrence of vectors; and “disease models” (DM) using, as dependent variables, the occurrence of disease cases in humans. Different pathogeographic scenarios led to DMs that differed in terms of the predictor variables included (see details in section 3.2). VMs and DMs were combined as shown in Table 1, in order to obtain different TMs. Finally, the resulting TMs were compared through the assessment of classification and discrimination capacities respect to the dataset of reported human cases used for training models. The chosen TM was the one that best fitted the distribution of human disease cases, leading to the most robust assumption on the limiting role of vectors and carriers for the LB zoonotic case.

**TABLE 1.** Method for building transmission model according to different pathogeographic scenarios, based on the combination of disease models (DM) and vector models (VM). Lower-case letters in parentheses represent sets of predictor variables included in the model, related to different factors: environment (e), and wild carriers (c). The vector factor, when used as a predictor variable within a DM (see CL and NL pathogeographic scenarios), is represented by the output of a VM (i.e., the degree of favourability for vectors to occur). The symbol “ꓵ” means “fuzzy intersection”. Details on the method for DM and VM building, and on the use of environmental and reservoir variables, are given in the main text.

| Pathogeographic scenario* | Transmission Model (TM) |
| --- | --- |
| VCL | $DM(e) \cap DM(c) \cap VM$ |
| CL | $DM(e, VM) \cap DM(c)$ |
| VL | $DM(e, c) \cap VM$ |
| NL | $DM(e, c, VM)$ |

\*VCL: both vectors and wild carriers are geographically limiting factors; CL: only carriers are limiting factors; VL: only vectors are limiting factors; NL: neither vectors nor carriers are limiting factors, i.e. vector and carriers are mutually compensating factors.

### 2.2 Study area

Our study area was established in the biogeographical context of the Holarctic region according to Udvardy (Fosberg FR. A, 1976). This area covers the historical distribution of Lyme borreliosis (LB) since the 1980s decade, when the first LB human cases were confirmed (Dennis & Hayes, 2002). Although *B. burgdorferi* s.l. has been detected in some countries of the southern hemisphere, there is currently no certain evidence of its presence in human beings (Beaman & Beaman, 2016; Chalada et al., 2016; Ozdenerol et al., 2015). The southern limits of the Holarctic region, far from consisting on crisp lines, are defined as biogeographical transition zones (Kreft & Jetz, 2013; Morrone, 2015). Compared to other global regionalizations (Cox, 2001; Holt et al., 2013; Procheş & Ramdhani, 2012; Wallace, 1876), Udvardy (Fosberg FR. A, 1976)’s includes in the Palearctic region areas considered to be transitional (Kreft & Jetz, 2013; Morrone, 2015). Thus, it extends the Saharan limit southward up to the Sahel; and includes the Himalayan highlands, the Szechwan highlands, and the Chinese subtropical forests. We also included Central America in our study area in order to take the Mexican transition zone (Kreft & Jetz, 2013; Morrone, 2005) into account. This area is considered to be of high importance for the diversification of mammal hosts in parallel with the diversification of their parasites (Morrone & Gutiérrez, 2005).

Due to the lack of georeferenced records of LB in humans (Blanchard et al., 2022), we used as spatial units the administrative divisions at the national level, and at sub-national levels for those countries with notably larger surface areas: Russia, Canada, China, and the United States of America.

### 2.3 Modelling technique

The outputs of our distribution models were expressed in terms favourability values (*F*), ranging from 0 to 1, which were calculated using the Favourability Function (Acevedo & Real, 2012; Real et al., 2006):

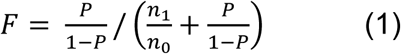

where *P* is the spatial probability of occurrence of the modelled entity, and *n_1_* and *n_0_* are the number of administrative units in which the modelled entity was present and absent, respectively. *P* values were calculated through a logistic regression:

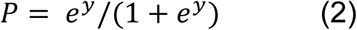

where *e* is the base of Napierian logarithms and *y* is the logit function (i.e., a linear combination of predictor variables). For this regression, the dependent variables were the presence/absence (1/0) of reports of LB in humans in DMs; and the presence/absence of vector records in VMs. Predictor variables in the model were selected through a conditional forward-backward stepwise procedure: variable entry was based on the signification of Rao’s (Rao, 1948) score tests, and variable elimination was based on likelihood ratio probabilities. Variable coefficients were estimated using a machine-learning iterative process based on the maximum log-likelihood criterion. These regressions were trained using IBM-SPSS Statistic v.28.

Preliminary controls were made for minimizing excessive multicollinearity and type-I error. Multicollinearity was controlled by preventing the coexistence of predictors with a Spearman correlation coefficient >0.8 in the same model. When this coexistence was detected, we eliminated the variable with the lowest Rao’s score value from the variable set proposed for model entry, and then trained the logistic regression again. Type-I error, which could arise from the use of a large amount of predictor variables, was avoided using the false discovery rate (FDR) (Benjamini & Hochberg, 1995). Thus, only variables with Rao’s *p* < *i*×*q*/*v* were proposed for entry in the model, where *i* is the position of the variable arranged in increasing *p* order, *q* = 0.05, and *v* is the total number of predictor variables initially proposed. Additionally, the use of a data set based on presence/absence in administrative units prevented autocorrelation caused by sample bias at the subnational level. Spatial autocorrelation due to natural factors, such as patterns linked to the evolutionary history (e.g., the geographical origin of the species, limits to dispersal), was considered through a trend surface approach (Legendre & Legendre, 1998) (see section 2.4).

Model performance was assessed using Hosmer and Lemeshow goodness of fit (2000). The significance of variables included in the models was evaluated according to Wald tests.

### 2.4 Vector models (VM)

We considered the main vectors of *B. burgdorferi* s.l., which are *Ixodes ricinus* and *I. persulcatus* in Eurasia, and *I. scapularis* and *I. pacificus* in America (Steere et al., 2005). We also included as potential vectors those *Ixodes* species involved in enzootic cycles, since they maintain the circulation of the *B. burgdorferi* complex in wildlife populations (Piesman & Gern, 2004). Furthermore, the pathogen has been isolated from several ixodid species, other than *Ixodes*, such as *Haemaphysalis*, *Dermacentor* and *Amblyomma* (specifically *Amblyomma americanum*) (Eisen & Lane, 2002). The vector capacity of non-*Ixodes* ticks is uncertain and currently discussed, as some authors have found them to be incompetent to transmit *B. burgdorferi* (Breuner et al., 2020). However, LB human cases were found in areas from where *Ixodes* species were absent (Elhelw et al., 2014); so, the presence of a different tick genus needs to be assumed. To account for the latter possibility, we built two alternative VMs that differed in terms of the tick species considered as dependent variables: (1) “*Ixodes*” VM, with only *Ixodes* species considered; and (2) “tick” VM, with species from the genera *Ixodes, Haemaphysalis*, *Dermacentor,* and *Amblyomma* assumed to be competent vectors. For that purpose, we selected those species whose vector capacity has been or might be confirmed, according to Eisen(Eisen, 2020): *I. scapularis*, *I. pacificus*, *I. spinipalpis*; *I. ricinus, I. persulcatus*, *I. sinensis*, *I. uriae, and I. angustus* for the *Ixodes*-VM; and the same ones plus *H. longicornis*, *H. concinna*, *D. andersoni*, *D. variabilis*, *D. occidentalis*, *D. nuttalli*, *D. silvarum*, and *A. americanum* for the Tick-VM. Information from tick distributions was obtained from the European Centre for Disease Prevention and Control (ECDC), from Centres for Disease Control and Prevention (CDC), and from the literature (Table S1). Tick presences/absences were projected to the national/sub-national level.

We built a favourability model for the distribution of each tick species, using the presence/absence of the species as dependent variable, and environmental predictor variables related to different factors: climate, topo-hydrography, human activity and presence, livestock, ecosystem types, and agriculture (Table S2). A trend surface variable (Legendre, 1993) was also included, whose values were calculated by running a favourability model only based on spatial predictors (i.e., monomials for latitude and longitude combinations up to the third-degree; see Legendre (1993), and Legendre *et al*. (1998)) that were backward-stepwise selected. The calculation of trend surface variables was contextualized in the continent where the species was present (i.e., Palearctic or Nearctic). So, in the continent from where a species was absent, the spatial favourability value was assumed to be 0. In the event that the spatial factor explained most of the variation in favourability by itself (i.e., either the variable selection included only the spatial variable, or the sum of Wald values for the other variables was <10% compared to that of the spatial variable), we made two different models for every species, which were combined using fuzzy logic. These models were: a model based on environmental predictors, and a model based solely on the spatial variable. Both models were then combined using the fuzzy intersection.

Finally, VMs were built as the combination of all the corresponding single-tick-species favourability models using the fuzzy union; that is, by retaining, for every administrative unit, the highest degree of membership (i.e., the maximum favourability value) recorded in all the models (Zadeh, 1965). This criterion responded to the assumption that a single vector species could imply potential for pathogen transmission.

### 2.5 Disease models (DM)

#### 2.5.1 The distribution of Lyme borreliosis in humans

The dependent variable, in all DMs, was the presence/absence of reports of LB in humans in national/sub-national units. Incidence data and reported cases, since the 1980s decade until 2022, were initially collected from the Global Infectious Disease and Epidemiology Online Network (GIDEON). In order to complete GIDEON’s information, a complementary literature search was made (Table S3). In the case of USA, the CDC informs on “low incidence states” that are not included in GIDEON. As LB georeferenced cases in the CDC databases were referred to the patient’s county of residence, it was impossible to distinguish autochthonous cases from imported ones. Thus, the presence of LB in low incidence states, according to CDC, was contrasted with the information provided by the Health Departments of the corresponding states.

#### 2.5.2 Wild carriers

We considered as wild carriers of *B. burgdorferi* s.l. small mammals from the orders Rodentia, Eulipotyphla and Lagomorpha. So, all potential pathogen carriers, confirmed and possible wild reservoirs, are included in the risk analyses. Although some species of lizards and birds are found to be reservoirs of LB (Steinbrink et al., 2022), they were not considered here due to the uncertainty about their contribution to large-scale biogeographical transmission of *Borrelia* genospecies infecting humans; and because some of these species seem to be dead-end hosts (Piesman & Gern, 2004). We included small mammal chorotypes, i.e., distributional pattern shared by a group of species (Real et al., 2008), as predictor variables representing the presence of wild carriers in our DMs. This approach was proposed by Olivero *et al*. (2017) as a way to integrate reservoirs in pathogeographical models when the knowledge of the reservoir species complex is imprecise. Chorotypes were defined for the whole set of species belonging to orders Rodentia, Lagomorphs and Eulipotyphla. For this purpose, we took the following steps: (1) Species distributions were provided by the IUCN and the Handbook of the Mammals of the World (Wilson et al., 2016, 2017); and projected into national and sub-national spatial units as a presence/absence matrix. *Mus musculus*, *Rattus rattus*, and *Rattus norvergicus* were excluded from the analyses due to their ubiquitous distribution might not affect the capacity of distinguishing favourable areas for LB transmission. (2) Distribution ranges were classified hierarchically according to the Baroni-Urbani & Buser’s similarity index (Baroni-Urbani & Buser, 1976), and to the unweighted pair-group method using arithmetic averages (UPGMA) (Sneath and Sokal 1973, Kreft and Jetz 2010). (3) Statistical significance of the resulting clusters was assessed using the RMacoqui 1.0 software (http://rmacoqui.r-forge.r-project.org/) (Olivero et al., 2011). Clusters representing significantly similar species distributions were considered as chorotypes.

Chorotypes were turned into predictor variables useful for the DMs through the chorotype species richness, meaning, the number of extant species forming part of the chorotype for every spatial unit. As with environmental predictors, an FDR test was used in order to avoid excessive type-I error when included these chorotypes in DMs.

#### 2.5.3 The environment

For including the environmental factor in DMs, we took into account the same environmental predictors as the tick species models (Table S2). In addition, DMs considered deer as amplifiers of the spatial risk of LB (van Wieren & Hofmeester, 2016). Thus, we included as predictor variables the conditions favouring the occurrence of deer in North America and Eurasia, that is, favourability values obtained from models trained on the presence/absence of deer species. These models were made following the same method and variables exposed above for ticks. The deer species considered were: *Alces alces*, *Capreolus* (*Ca.*) *capreolus*, *Ca. pygargus*, *Cervus* (*C.*) *albirostris*, *C. canadensis*, *C. elaphus*, *C. hanglu*, *C. nippon*, *Dama dama*, *D. mesopotamica*, *Hydropotes inermis*, *Muntiacus reevesi*, *Odocoileus hemionus*, *O. virginianus* and *Rangifer tarandus*. Native distributions were obtained from the IUCN, corrected with the Handbook of the Mammals of the World (Mattioli, 2011), and projected into national/sub-national spatial units. We also considered the introduced population of *D. dama, M. reevesi, H. inermis, O. virginianus,* and *C. nippon,* whose distributions were also obtained from the UICN and contrasted with the literature (Table S4). When a species had native and introduced populations, the distribution of both were modelled separately, as the presence of introduced deer is not attributable to their evolutionary history but to the human activity. These two models were later assembled using the fuzzy union (i.e., by retaining the maximum favourability value recorded in the two models). So, the role of deer on the *B. burgdorferi* s.l. circulation in the wild was constrained by their occurrence areas, favourability values for a species were multiplied by the species presence/absence (1/0).

#### 2.5.4 Disease models (DMs) according to different predictor factors

We finally built five DMs describing favourable areas for the distribution of LB cases in humans, based on different sets of predictor variables:

- DM(e): only environmental predictors.
- DM(c): only wild carrier chorotypes used as predictors.
- DM(e,c): both environmental predictors and wild carrier chorotypes were introduced in “blocks”; i.e., at a first stage only environmental predictors were included in the stepwise variable selection. Chorotypes were then added along a second stepwise selection stage.
- DM(e,VM): environmental predictors were stepwise selected together with the favourability for the presence of vectors (i.e., with the favourability output of the VM).
- DM(e,c,VM): environmental predictors were stepwise selected together with the favourability for the presence of vectors and with wild carrier chorotypes, the latter along a second stepwise selection stage.

### 2.6 Transmission models (TM)

The combination of four pathogeographical scenarios (see Table 1) and two alternative VMs (*Ixodes*-VM and tick-VM) resulted in a total of eight TMs.

### 2.7 Model selection and downscaling

Model performance was assessed using Hosmer and Lemeshow goodness of fit (2000). The significance of variables included in the models was evaluated according to Wald tests. Model discrimination capacity was assessed according to the area under the “receiver operating characteristic” curve (AUC) (Lobo et al., 2008). Classification capacity was assessed according to the 0.5 favourability threshold, a statistically relevant value at which the probability of presence is equal to the overall prevalence (Acevedo & Real, 2012). The indices applied for classification assessment were sensitivity, specificity, the Correct Classification Rate (CCR) (ranging from 0 to 1); True Statistic Skill (TSS) (ranging from -1 to 1); over-prediction and under-prediction rate (ranging from 0 to 1) (Barbosa & Real, 2012; Fielding & Bell, 1997).

A transmission model was finally selected after comparisons according to their discrimination and classification capacities, in order to select the most fitted TM respect to the distribution of disease cases. The above-mentioned indices were given different relevance along the model selection procedure. When comparing TMs built considering different zoonotic scenarios but a same VM, we gave preference to indices based on balanced information regarding the presence and the absence of LB cases recorded, such as the AUC, the TSS and the CCR. We did it because the surface area predicted to be at risk of transmission is expected to decrease with the number of geographically limiting factors considered. The sensitivity of a TM is the proportion of countries with LB cases recorded that are identified as high-risk areas by the model, whereas specificity is the proportion of countries without LB cases recorded that are identified as low-risk areas (the threshold being F=0.5). Hence, models based on the VCL scenario should be expected to be, *a priori*, less sensitive but more specific than models based on the NL scenario. In addition, underprediction is the proportion of low-risk areas (according to the model) corresponding to countries with LB cases recorded, whereas overprediction is the proportion of high-risk areas corresponding to countries without LB cases recorded. So, the VCL scenario should be expected to show, *a priori*, higher underprediction but lower overprediction rates than the NL scenario.

Instead, when comparing TMs built considering different VMs but a same zoonotic scenario, we gave preference to sensitivity, specificity, underprediction and overprediction. For conservative purposes regarding predictions, considering that the object of study was the transmission risk of a disease, higher sensitivity was preferred to higher specificity, and lower underprediction was preferred to lower overprediction.

The TM selected was finally projected into a grid of 11,486 hexagons of 7,774-km^2^ (https://zenodo.org/records/10028166), using a downscaling approach (Bombi & D’Amen, 2012). The downscaling consisted of projecting the favourability into a set of variables considered in the hexagon grid, for which we used the following favourability (*F*) equation (Real et al., 2006):

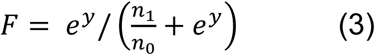

where *y* is the same logit function as in equation 2 (i.e., a linear combination of predictor variables), and *n_1_* and *n_0_* are the number of administrative units in which the modelled entity was present and absent, respectively.

### 2.8 Assessment of predictive capacity of the Transmission Model

In order to evaluate the predictive potential of the downscaled TM, we assessed its discrimination and classification capacities respect to two countries with entirely different biogeographical contexts, USA and China, for which high-resolution georeferenced information on LB cases is available in the whole territory. The assessment was performed separately for both countries. For USA, we used the database of georeferenced cases by county collected by the CDC in 2017, when the highest number of yearly cases in that country was reported (CDC, 2022). For China, we used a database of reported cases provided by Che *et al*. (2022).

## 3. RESULTS

### 3.1 Vector Models‒VM

Relevant environmental predictors in most individual vector models were temperate forests and shrublands. Extreme climatic environments, such as arid ecosystems and tundra, were significant variables for *I. ricinus* and *I. uriae*, respectively (Table S5). Besides, climate predictors such as temperature annual range and precipitation seasonality seem to explain favourable conditions for the distribution of *Haemaphysalis, Dermacentor*, and Eurasian *Ixodes* ticks. Among the anthropogenic predictors based on human activity, livestock was positively related to the presence of *A. americanum*, *D. occidentalis*, and the Eurasian population of *H. longicornis*; and agriculture showed a negative relation with the presence of *Haemaphysalis* and *Ixodes* species. The closeness of human-populated areas also seemed to favour the distribution of *I. ricinus*. Finally, slope and elevation described favourable conditions for the presence of American tick species, being the most significant variables in the models for the American *Ixodes* species, for *A. americanum*, and for *D. andersoni*.

The resulting *Ixodes-*VM shows favourable conditions (F > 0.2) for the presence of *Ixodes* vectors throughout North America and Europe, and in most of northern, central, and eastern Asia excluding South Korea (see Figure S1). In Africa, environmental conditions seem to be favourable to the presence of *Ixodes* ticks only in Morocco and Tunisia. High favourability areas (F > 0.8) are, however, concentrated in the eastern and western territories of USA and Canada, in central and northern Europe, in China, Mongolia and Japan, and in large extensions of Siberia. Compared to the *Ixodes-*VM, the Tick-VM expands the high favourability areas to the centre and south of North America, and to most of the Asian continent, including South Korea (Figure S1).

### 3.2 Disease models‒DM

#### 3.2.1 Disease model based on carriers‒DM(c)

The biogeographical analysis of the distribution of 1,169 potential carrier species led to the definition of 74 chorotypes, of which 16 ones overcame the FDR test and were thus proposed as predictors in disease models (see Figure S2). In the DM(c), chorotypes 19, 30, 48 and 56 contributed significantly to explain the presence of LB cases in humans (F > 0.2) in Mexico and the eastern half of North America, in Europe, and in most of the Asian continent, only excluding Japan, South Korea, South-eastern Asia (Table S6 and Figure S3).

#### 3.2.2 Disease model based on the environment‒DM(e)

Favourable conditions for the occurrence of LB cases were explained by climate predictors (low minimum temperatures of the coldest month); topography (low slopes); human activity predictors (closeness to railways, cropland mosaics); forest (broadleaved deciduous and needleleaved evergreen forests); and the presence of favourable conditions for deer species: the Siberian roe deer (*Ca. pygargus*), fallow deer (*D. dama*), and Reeves’ muntjac (*M. reevesi*) (Table S6). The DM(e) provides a similar picture to the DM(c) on the distribution of favourable areas for the presence of LB cases in Eurasia, but the former contributes better to explain the presence of cases in western North America, China, South Korea, and Japan; while excluding Mexico as a favourable area (Figure S3).

#### 3.2.3 Disease model based on both the environment and carriers‒DM(e,c)

When carriers were considered as predictors together with the environment, chorotypes 19, 30, 44 and 56 were included in the model, along with the variables included in the DM(e) (Table S6). Compared to the DM(e), the DM(e,c) was able to predict the occurrence of LB cases in northern Africa, specifically in Algeria and Egypt (Figure S3).

#### 3.2.4 Disease model based on both the environment and vectors‒DM(e,VM)

The DM(e,VM), when the *Ixodes-*VM was employed, included climate (high temperature annual range); human activity (closeness to railways); forest (needleleaved deciduous forests); and favourable conditions for the Siberian roe deer (*Ca. pygargus*), the red deer (*C. elaphus*), and the moose (*A. alces*). When using the Tick-VM, cropland mosaics and mixed broadleaved and needleleaved forests were additionally included; and favourable conditions for the presence of deer were focused on the Siberian roe deer (*Ca. pygargus*) and the Eurasian fallow deer (*D. dama*) (Table S6). Areas favourable to the occurrence of LB cases were similar to those described by the DM(e,c), except in northern Africa. There, with the *Ixodes-*VM, only Morocco and Tunisia were considered favourable areas (F>0,2) for the presence of LB cases; with the Tick-VM, northern Africa was excluded from the favourable zone (Figure S3).

#### 3.2.5 Disease model based on the environment, carriers, and vectors‒ DM(e,c,VM)

The two DMs(e,c,VM) included the same environmental predictors as their corresponding DMs(e,VM) (Table S6). Both DMs(e,c,VM) pointed to Morocco, Algeria, and Tunisia as north-African countries favourable to the occurrence of LB cases (Figure S3).

### 3.3 Transmission models‒TM

TM maps are shown in Figure S4. The two TMs resulting from the VCL scenario (i.e., both vectors and carriers are geographically limiting factors) were geographically very restrictive to the consideration of transmission risk. Thus, regions such as western North America, Mexico, northern Africa, Japan, South Korea, and southern China were excluded from risk areas despite having a record of LB cases (Figure S4). Also, high risk (F>0,8) was only noted in eastern USA, central Europe, and north-eastern Asia. This picture is similar to that offered by the two TMs resulting from the CL scenario (i.e., only wild carriers are geographically limiting factors). The only relevant difference was an increase in surface of the high-risk area (F > 0,8) in eastern Europe and in eastern USA. In contrast, the TMs resulting from the VL scenario (i.e., only vectors are geographically limiting factors) added three significant regions with recorded cases of LB to the risk area: western North America, Japan, and southern China (Figure S4). Finally, northern Africa was also considered to be inside the risk area by the two TMs based on the NL scenario (i.e., neither vectors nor carriers are geographically limiting factors), which are described above as the DMs(e,c,VM).

### 3.4 Model assessment and final transmission model selection

#### 3.4.1 Step-model assessment

The AUC values of all the models made as steps for building transmission models were >0.8, pointing to excellent discrimination capacity according to Hosmer and Lemeshow (2000). Deer and tick distribution models presented average TSS values of 0.84 (SD=0.145) and 0.79 (SD=0.115) respectively, and CCR values >0.8; with the exception of the model of *C. nippon*, whose TSS value was 0.69. Regarding VMs, average TSS values were 0.49 (SD=0.002) and average CCR values were 0.81 (SD=0.023). DMs showed an average TSS of 0.70 (SD=0.032) and an average CCR of 0.86 (SD=0.018). The values for all indices regarding each tick and deer species model are shown in Table S7; and values for VMs and DMs are shown in Table S8.

#### 3.4.2 Transmission model (TM) assessment and selection

The eight resulting TMs presented AUC values higher than 0.91, pointing to outstanding discrimination capacities according to Hosmer and Lemeshow (2000) (see Table 2). In all the models, TSS values were higher than 0.66 and CCR values were higher than 0.8. Sensitivity and specificity values were always higher than 0.73 and 0.85, respectively; while underprediction and overprediction rates were always lower than 0.35 and 0.08, respectively (see Table 2).

**TABLE 2.** Transmission Models (TM) assessment based on discrimination and classification capacities respect to the presence/absence of reported Lyme borreliosis cases. AUC: area under the receiver operator characteristic curve; FCT: favourability classification threshold; TSS: True Skill Statistic; Sens.: sensitivity; Spec.: specificity; CCR: correct classification rate; Underp.: underprediction rate; Overp.: overprediction rate.

| Transmission Model |  | AUC | FTC | TSS | Sens. | Spec. | CCR | Underp. | Overp. |
| --- | --- | --- | --- | --- | --- | --- | --- | --- | --- |
| With<br>*Ixodes*-VM | TM(VCL*) | 0.922 | 0.5 | 0.685 | 0.732 | 0.952 | 0.809 | 0.346 | 0.033 |
|  | TM(VL*) | 0.946 | 0.5 | 0.778 | 0.874 | 0.905 | 0.884 | 0.208 | 0.055 |
|  | TM(CL*) | 0.921 | 0.5 | 0.667 | 0.763 | 0.905 | 0.812 | 0.331 | 0.062 |
|  | TM(NL*) | 0.948 | 0.5 | 0.761 | 0.904 | 0.857 | 0.888 | 0.174 | 0.077 |
| With<br>Tick-VM | TM(VCL*) | 0.915 | 0.5 | 0.671 | 0.747 | 0.924 | 0.809 | 0.340 | 0.051 |
|  | TM(VL*) | 0.942 | 0.5 | 0.756 | 0.889 | 0.867 | 0.881 | 0.195 | 0.074 |
|  | TM(CL*) | 0.926 | 0.5 | 0.688 | 0.793 | 0.895 | 0.828 | 0.304 | 0.065 |
|  | TM(NL*) | 0.950 | 0.5 | 0.761 | 0.894 | 0.867 | 0.884 | 0.188 | 0.073 |

\* VCL: both vectors and carriers are geographically limiting factors; CL: only carriers are assumed to be geographically limiting factors; VL: only vectors are assumed to be limiting factors; NL: neither vectors nor carriers are assumed to be limiting factors.

In the TM selection process, we first selected those ones resulting from the NL scenario because they showed the highest AUC values (>0.94) and the highest CCR values (>0.88). Besides, the NL scenario also provided the highest TSS value among the TMs using the Tick-VM, and the second highest TSS value among the TMs using the *Ixodes*-VM. In both cases, the TSS value was 0.761. Then, between the two TMs based on the NL scenario, we selected that one using the *Ixodes*-VM because this option provided higher sensitivity and lower underprediction than the TM using the Tick-VM.

### 3.5 Downscaled Lyme borreliosis transmission model

The selected transmission model provided details in the perception of geographical areas at risk of LB transmission that are hardly seen in the original output, while they are consistent with the LB case record (compare Figure 2-B). Thus, in America, downscaled risk areas (F > 0.2) were predicted in Mexico; in Africa, the Egyptian lands adjacent to the Nile River joined to the Maghreb countries as risk areas; in Asia, some scattered risk locations are predicted in South Korea. Besides, the downscaled model avoided designating risk areas (F > 0.2) and high-risk areas (F > 0.8) in extreme latitudes within some countries at risk according to the original model. This is the case of British Columbia (Canada) and of some federal subjects of northern Siberia (specially the Republic of Sakha), where either risk or high-risk areas were restricted to the south; of Mongolia, Kazakhstan, western China and some Mediterranean peninsulas (Spain, Portugal, Italy), where either risk or high-risk areas were restricted to the north; and of the Maghreb in North Africa, showing risk only in its median latitudes. Finally, the intermediate risk (0.2 > F > 0.8) predicted in Eastern-Siberia (specifically in the Republic of Sakha, the Magadan Oblast, and the Chukotka Autonomous Okrug) should not be taken into account, as the model downscaling were not reliable in this zone. Climate predictors showed, in this area, rescaled values outside the range in which the models were calibrated (see Figure 1-E).

**FIGURE 1.**
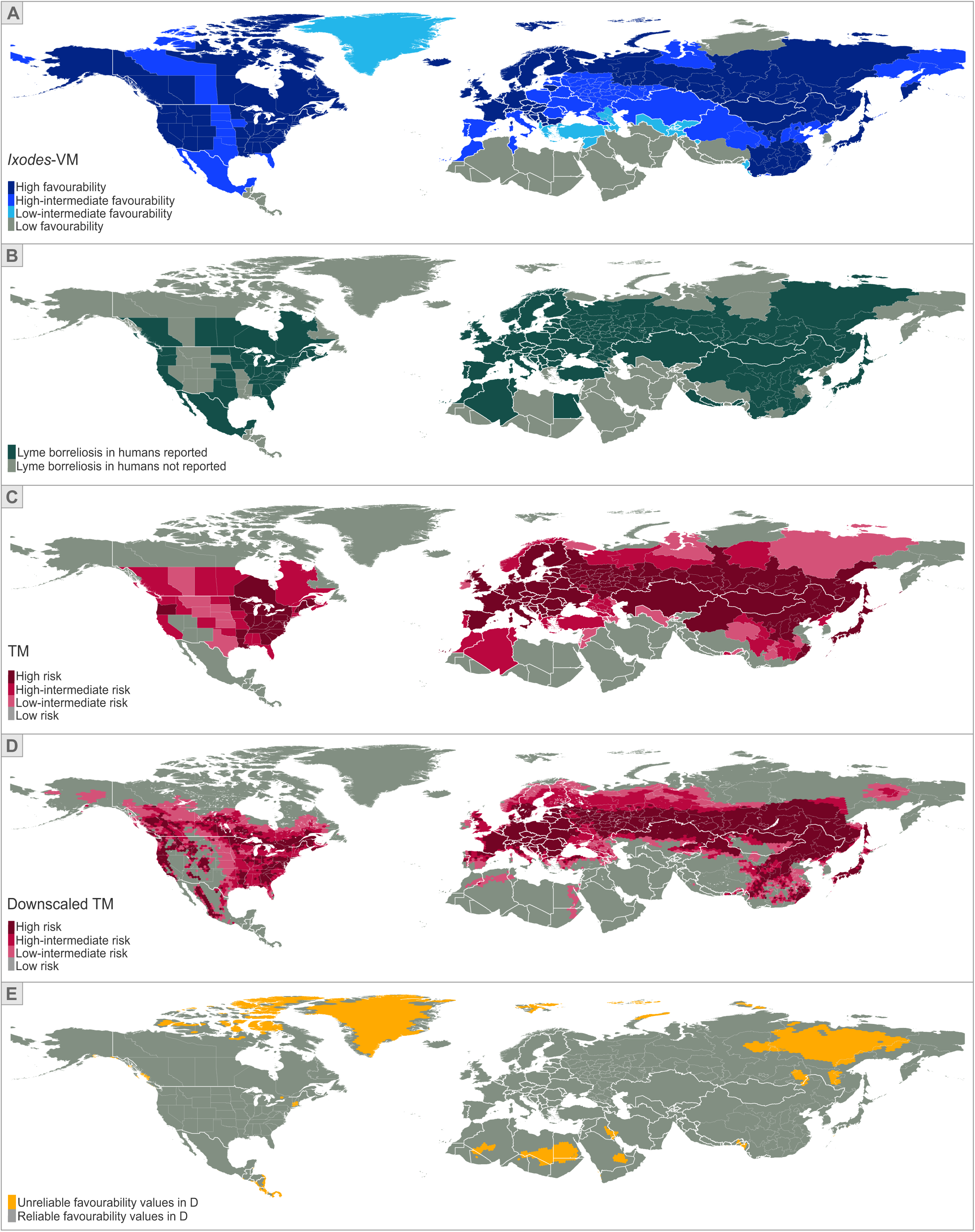
Relevant maps driving to the Lyme borreliosis transmission risk model. **(A)** *Ixodes* vector model (VM), resulting from the fuzzy union of favourability models of tick species belonging to genus *Ixodes*; favourability was based on environmental and human predictors. (**B)** Distribution of Lyme borreliosis in humans, used as the dependent variable for disease model calibration. (**C)** Lyme borreliosis transmission risk model (TM, based on environmental variables, human predictors, carrier chorotypes and the VM) at the original spatial resolution (national/subnational administrative units). **(D)** Lyme borreliosis transmission risk model (TM) downscaled to a 7.774-km^2^ resolution. (**E)** Zones where favourability values in A are unreliable, as model predictors at the downscaled resolution show values beyond their range at the calibration resolution. Risk and favourability values were categorized as low (<0.2), low-intermediate (0.2-0.5), high-intermediate (0.5-0.8), and high (>0.8).

**FIGURE 2.**
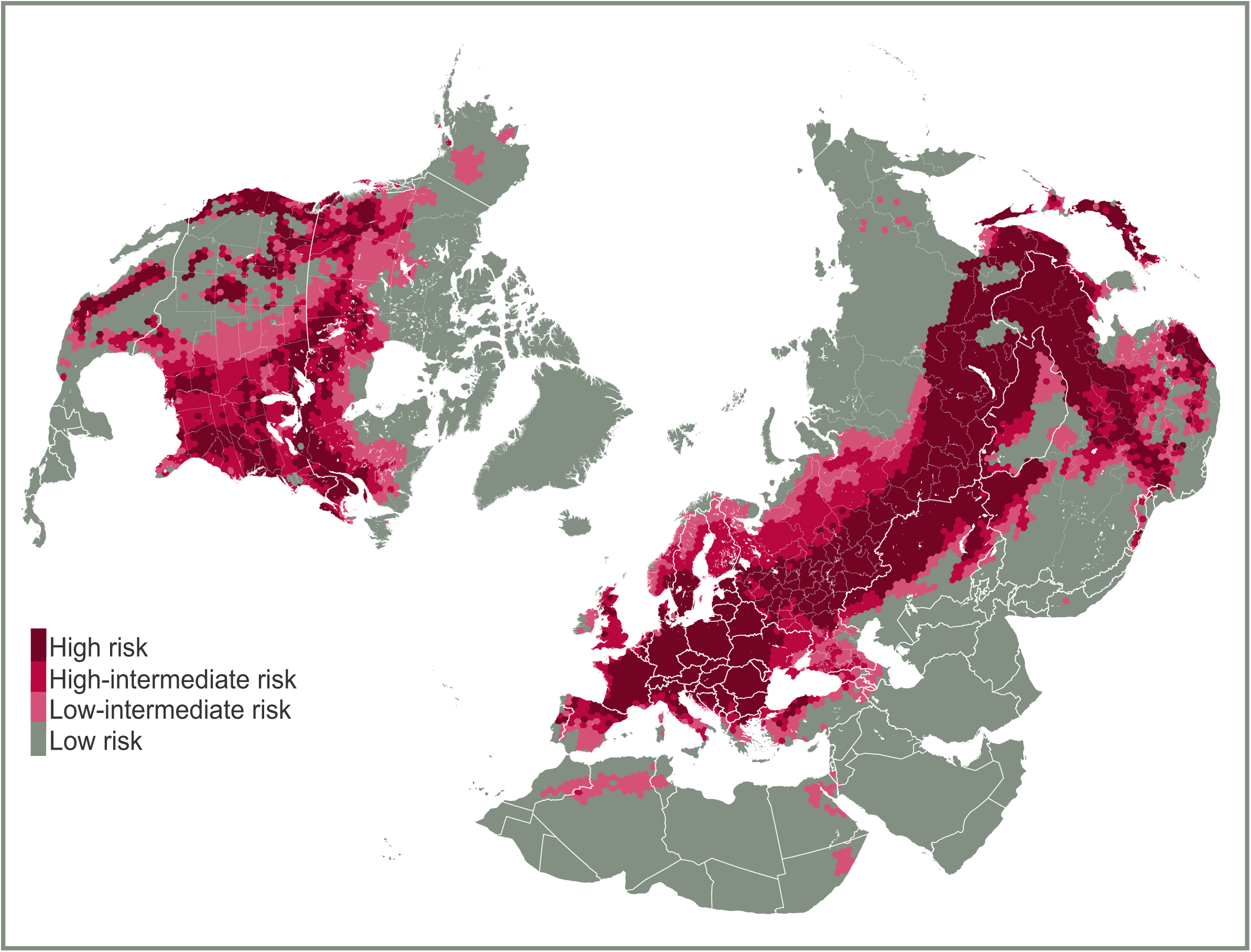
Lyme borreliosis transmission risk map (spatial resolution: 7,774-km^2^ hexagons). Risk values were categorized as low (<0.2), low-intermediate (0.2-0.5), high-intermediate (0.5-0.8), and high (>0.8). This model corrects the one in Figure 1-D considering the map of unreliable areas shown in Figure 1-E.

### 3.6 Assessment of predictive capacity of the Transmission Model

AUC values for predictive capacity assessments of the downscaled TM, in USA and China, were 0.764 and 0.675, respectively, pointing to good discrimination capacity according to Hosmer and Lemeshow (2000). Regarding classification capacities, in USA, the TSS was 0.47 and the CCR was 0.71. In China, the TSS was 0.25 and the CCR was 0.59. Underprediction was 0.094 (SD=0.009) in both countries, whereas overprediction was 0.656 (SD=0.141). The values for all the assessment indices are shown in Table S9.

## 4. DISCUSSION

### 4.1 Geographical risk pointing out a possible future expansion into northern latitudes

We have built the first global Lyme borreliosis risk model at a subnational resolution, considering the most comprehensive distribution dataset of reported human cases to date, and the combined biogeography of vectors and wild carrier hosts. Our results confirmed risk of LB transmission in endemic areas of North America, Eurasia and North Africa. Implementing a downscaled approach enabled us to locate risk areas with an unprecedented spatial resolution, overcoming the insufficient epidemiological information available at the subnational level.

The lack of effective surveillance hinders the validation of predictive power of our model in regions such as northern Africa, where the analysis of tick-borne disease risk is virtually non-existent (Perveen et al., 2021). In contrast, the availability of information in USA and China suggested that our model was able to make valid predictions at spatial resolutions finer than the national scale, and even beyond well surveyed areas. For example, in USA, the model predicted high risk in some areas where the presence of LB cases was reported in ambiguous terms (e.g., where the autochthonous nature of the cases was not warrantied), consequently counting as “absences” in the data set we used for model calibration. This happened particularly in scattered locations of heavily forested areas of the Grand Canyon and the Rocky Mountains, surrounded with large non-endemic territories. On the other hand, a visual inspection in China shows a scattered pattern of risk that aligns with the model published by Che et al.(2022).

The predictive assessment of the risk distribution for the rest of the study area is hampered by the lack of high-resolution epidemiological data. Our transmission model highlights a large risk area in Northern Eurasia, covering from Scotland and Norway in the West to Khabarovsk Krai (Russia) in the East. High-risk areas extend as far as the 60th parallel (see Figure 2). This boundary is recognized by some authors as the northern limit for tick-borne diseases (Rizzoli et al., 2011). However, according to our model, some level of risk of LB transmission extends northwards in Europe and West Siberia beyond endemic areas. Tick abundances could be experiencing an increase in the Nordic countries that is attributed to mild winters caused by the global climate change (Medlock et al., 2013). This situation results in an increased deer abundance and their migration northwards and towards higher altitudes (Jaenson et al., 2012). The incidence of tick-borne diseases in humans, such as the tick-borne encephalitis, has been increasing in the region over the last few decades (Van Heuverswyn et al., 2023).

Likewise, we have found a scenario akin to this in Canada. The distribution of high-risk areas (F > 0.8), predicted to be mostly located in the south-eastern provinces by the downscaled model, is consistent with the concentration of LB cases next to the borders with USA (Gasmi et al., 2022). Northwards, the predicted trend is just a decreasing gradient of intermediate risk values. In recent years, the expansion of *I. scapularis* ticks into the Canadian Prairie and Central regions has been well-documented (Clow et al., 2017). Thus, the spread of ticks into non-endemic areas, through dispersal linked to deer migration, could eventually translate into a LB spread. In the same geographical context, the Canadian health authorities have analysed the LB risk distribution relying solely in the established populations of *I. scapularis* and *I. pacificus* (Public Health Agency of Canada, 2022). This has led them to a more conservative view compared with ours, according to which the risk is restricted to the borders with USA. Instead, we propose that all possible factors potentially favouring the spread of the pathogen, such as alternative potential vectors, be considered.

In contrast, in southern Europe, the downscaled model provided us with a more conservative risk distribution, limited to the northern regions of the Mediterranean peninsulas. The drier Mediterranean climate results in less favourable conditions for high tick abundance (Medlock et al., 2013), so reducing transmission risk, as suggested by our vector models. The predictions of our transmission risk model are in line with the occurrence of cases reported in Italy in the last decades (Trevisan et al., 2023), and with the observation of a higher LB hospitalization rate in northern Spain (Amores Alguacil et al., 2022). Even more, the predicted risk in non-endemic countries such as Greece supports the assumption of undetected LB circulation in human populations (Karageorgou et al., 2022).

### 4.2 The role of vectors and carriers’ diversity in risk distribution

Our findings suggest that, although ticks and wild carriers are ecological factors needed for *Borrelia burgdorferi* s.l. transmission to humans (Piesman & Gern, 2004), both types of zoonotic agents could not be strictly limiting its occurrence at the biogeographical scale. That is, probably, because of the availability of potential vectors and reservoirs elsewhere throughout the study area. Instead, in spatial terms, these agents could be contributing to modulate the level of risk of LB transmission. The chances for pathogen transfer to humans would come out from the interaction of economic and leisure activities within environments where conditions foster the abundance of competent ticks and reservoirs; and probably, the contact between humans and vectors.

Moreover, our results support the role of *Ixodes* species as primary vectors for LB, but do not exclude other species to participate in the LB transmission. The first reason for this lies in our best-fitted models, which were provided by a pathogeographical scenario based on *Ixodes* species, but according to which these ones are not geographically limiting factors. Given that *Ixodes* species are not reported in all countries with LB cases, other ixodid ticks may be contributing to LB transmission. For instance, in Egypt (Elhelw et al., 2014) and in some South-Asian countries (Ji et al., 2022), our model predicted risk to be medium but not null. The second reason is that the transmission model built on the basis of four tick genera got good assessment values as well (see Table 1).

An essential key in the circulation of *Borrelia burgdorferi* s.l., and in the maintenance of tick populations, is the diversity of hosts in the environment. According to our model, in most of the study area, the presence of micromammals seems to influence the distribution of LB transmission risk beyond the environmental descriptors. The carrier chorotypes selected by the model, especially chorotypes SR30 and SR56, involve the presence of confirmed wild reservoirs such as *Peromyscus*, *Apodemus* and *Sorex* species. The community of carrier species, whether they are reservoirs or not, can influence the density of infected ticks which, in turn, affects the level of transmission risk (Kilpatrick et al., 2017). A high diversity of reservoirs, non-reservoirs carriers and “dead end” hosts results into a constant circulation of *Borrelia* species between ticks and mammals; in spite of which the effect of a healthy community of mammals may imply the decrease of risk through the “dilution effect” (Wood et al., 2014). This dilution consists of a decrease of zoonotic risk resulting from the local coexistence of both competent and non-competent pathogen carriers. In this context, disease control policies should be always considered in conjunction with wildlife conservation measures.

The human disturbance of natural environments (i.e., habitat fragmentation, landscape changes, forest conservation policies) could entail changes in the feed-net of ticks, leading to modifications in the level of LB transmission risk (Wood & Lafferty, 2013). Human impacts driving to defaunation in the natural ecosystems can derive in the increase of disease prevalence, resulting from the reduced predation on micromammals and, specially, on infected hosts (Levi et al., 2016; Young et al., 2014). For this reason, eradication policies focused on carriers and other hosts should be discarded, as their effects on disease prevalence can be unforeseen or even contrary to those expected (Amman et al., 2014; Rand et al., 2004).

## DATA AVAILABILITY

All data needed to evaluate the conclusions in the paper are present in the main text or the supplementary materials.

## Data Availability

All data produced in the present work are contained in the manuscript and the supporting information

## ACKNOWLEDGEMENTS

Funding statement: MC-M was supported in parts by the European Social Fund, Youth Guarantee contract (University of Malaga), with ID SNGJ5Y6-47, and the Project of the Spanish Ministry of Science and Innovation and European Regional Development Fund (ERDF) [PID2021-124063OB-I00]; AA-S was supported in parts by the II Plan Propio de Investigación, Transferencia y Divulgación Científica of the University of Malaga, postdoctoral contract, and by MOPGA 2024: Visiting Fellowship Program for early career researchers of the French Ministry for Europe and Foreign Affairs, in collaboration with the French Ministry for Higher Education and Research, and implemented by Campus France, France.

## Authors contribution

M.C-M: Conceptualization (supporting), Data curation (lead), Formal analysis (lead), Investigation (lead), Methodology (lead), Visualization (lead), Writing-Original Draft Preparation (lead), Writing-Review & Editing (lead). A.M-T: Conceptualization (supporting), Data curation (supporting), Formal analysis (supporting), Investigation (supporting), Writing-Review & Editing (supporting). A.A-S: Writing-Original Draft Preparation (supporting), Writing-Review & Editing (supporting). M.S: Investigation (supporting), Writing-Review & Editing (supporting). J.O: Conceptualization (lead), Formal Analysis (lead), Funding Acquisition (lead), Investigation (lead), Methodology (lead), Supervision (lead), Validation (lead), Visualization (lead), Writing-Original Draft Preparation (lead), Writing-Review & Editing (lead).

