## Supporting information for "Global risk assessment of Lyme borreliosis transmission"

[1] Grupo de Biogeografía, Diversidad y Conservación, Departamento de Biología Animal, Facultad de Ciencias, Universidad de Málaga, Malaga, Spain.[2] Maladies Infectieuses et Vecteurs: Ecologie, Génétique, Evolution et Contrôle (UMR MIVEGEC), Institut de Recherche pour le Développement (IRD), Institut national de recherche pour l'agriculture, l'alimentation et l'environnement (INRAE), Université de Montpellier (UM), Centre National de la Recherche Scientifique (CNRS), Montpellier, France.[3] Servicio de Sanidad Exterior, Centro de Vacunación Internacional, Ministerio de Sanidad, Consumo y Bienestar Social, Estación Marítima, Malaga, Spain.[4] Instituto IBYDA, Centro de Experimentación Grice-Hutchinson, Malaga, Spain.

### Table of Contents

**Table S1.** Literature and sources used to georeference the distribution of tick vectors.

**Table S2.** Independent predictor variables considered for distribution modelling.

**Table S3.** Literature used to georeference the distribution of reports of Lyme borreliosis in humans.

**Table S4.** Literature used to georeference the introduced free-living population of deer in Europe and North America.

**Table S5.** Vector species models' logit functions (i.e., linear combinations of predictor variables in the probability equations obtained with logistic regression).

**Table S6.** Disease model's logit functions (i.e., linear combinations of predictor variables in the probability equations obtained with logistic-regression).

**Table S7.** Assessment of tick and deer species models based on discrimination and classification capacities.

**Table S8.** Assessment of vector models (VM) and disease models (DM).

**Table S9.** Assessment of predictive capacity of the downscaled Lyme borreliosis transmission model.

**Table S10.** Deer model's logit functions (i.e., linear combinations of predictor variables in the probability equations obtained with logistic regression).

**Figure S1.** Tick vector distributions and favourability models.

**Figure S2.** Chorotypes considered for entry in disease models after overcoming a false discovery rate test.

**Figure S3.** Lyme borreliosis models.

**Figure S4.** Lyme borreliosis transmission risk models according to different pathogeographic scenarios.

**Figure S5.** Favourability models of tick species and their distribution within the study area.

**Figure S6.** Favourability models of deer species and their distribution within the study area.

**REFERENCES**

**Table S1. Literature and sources used to georeference the distribution of tick vectors.**

| | |
|---|---|
| ***Ixodes* species** | |
| *I. ricinus* | (ECDC, 2022; Kholodilov et al., 2021; Sultankulova et al., 2022) |
| *I. persulcatus* | (Černý et al., 2019; Dobler et al., 2019; ECDC, 2022; Inokuma et al., 2007; John et al., 2021; Mukhacheva & Kovalev, 2014; Popov & Popova, 2020; Popov & Yasyukevich, 2014; Rar et al., 2010; Sato et al., 2021; Turebekov et al., 2019; Zamoto-Niikura et al., 2018; Zhang et al., 2019) |
| *I. scapularis* | (CDC, 2022b; Eisen et al., 2016; Guzmán-Cornejo & Robbins, 2010; Kotchi et al., 2021; Scott et al., 2018) |
| *I. pacificus* | (CDC, 2022b; Eisen et al., 2016; Guzmán-Cornejo & Robbins, 2010; Kanji et al., 2022) |
| *I. angustus* | (Guzmán-Cornejo & Robbins, 2010; Hahn et al., 2020; Hutcheson et al., 2021; Lindquist et al., 2016; Pukhovskaya et al., 2019; Yamauchi et al., 2013; Yamborko et al., 2015) |
| *I. spinipalpis* | (Guzmán-Cornejo & Robbins, 2010; Hutcheson et al., 2021; Lindquist et al., 2016) |
| *I. uriae* | (Dietrich et al., 2011; Hahn et al., 2020; Kim et al., 2017; Lindquist et al., 2016; Muñoz et al., 2015; Munro et al., 2019; Sormunen et al., 2022) |
| *I. sinensis* | (Zhang et al., 2019) |
| ***Haemaphysalis* species** | |
| *H. concinna* | (Rubel et al., 2018; Zhang et al., 2019) |
| *H. longicornis* | (Kang et al., 2016; Kunwar et al., 2022; USDA National, 2023; Zhang et al., 2019; Zhao et al., 2020) |
| ***Dermacentor* species** | |
| *D. andersoni* | (Guzmán-Cornejo et al., 2016; Hutcheson et al., 2021; Lindquist et al., 2016) |
| *D. silvarum* | (Černý et al., 2019; Pukhovskaya et al., 2019; Rubel et al., 2020; Zhang et al., 2019) |
| *D. occidentalis* | (Guzmán-Cornejo et al., 2016; Paddock et al., 2018) |
| *D. nuttalli* | (Černý et al., 2019; Kulakova et al., 2014; Zhang et al., 2019) |
| *D. variabilis* | (CDC, 2022a; Guzmán-Cornejo et al., 2016; Hutcheson et al., 2021; Lindquist et al., 2016) |
| ***Amblyomma* americanum** | |
| *A. americanum* | (CDC, 2022c; Guzmán-Cornejo et al., 2011; Lindquist et al., 2016; Saleh et al., 2021; Springer et al., 2014) |

### Table S2. Independent predictor variables considered for distribution modelling.

| Factor | Variables | Code | Source |
|---|---|---|---|
| Climate | Annual mean temperature | *Bio1* | CHELSA (Karger et al., 2021) |
| | Maximum temperature of warmest month | *Bio5* | |
| | Minimum temperature of coldest month | *Bio6* | |
| | Temperature annual range (Bio5-Bio6) | *Bio7* | |
| | Annual precipitation | *Bio12* | |
| | Precipitation seasonality | *Bio15* | |
| Human concentration | Population Density | *Pop_den* | FAO (Salvatore et al., 2005) |
| | Distance to populated places | *Dis_pop* | FAO (Food and Agriculture Organization of the United Nations, 2005) |
| Infrastructures | Distance to roads | *Dis_road* | Harvard University (Harvard University, 2002) |
| | Distance to railways | *Dis_rail* | |
| Livestock | Density of buffaloes | *Buffaloes* | FAO (Food and Agriculture Organization of the United Nations, 2011) |
| | Density of cattle | *Cattle* | |
| | Density of goats | *Goats* | |
| | Density of horses | *Horses* | |
| | Density of pigs | *Pigs* | |
| | Density of sheep | *Sheep* | |
| Agriculture | Irrigated croplands | *Class11* | FAO (Food and Agriculture Organization of the United Nations, 2009) |
| | Rainfed croplands | *Class14* | |
| | Mosaic Cropland (>50) / Vegetation (<50%) | *Class20* | |
| | Mosaic Vegetation (>50%) / Cropland (<50%) | *Class30* | |
| | Percentage of areas equipped for irrigation | *AEI* | FAO (Food and Agriculture Organization of the United Nations, 2000) |
| Ecosystem types | Broadleaved evergreen and/or semi-deciduous forest | *Class40* | FAO (Food and Agriculture Organization of the United Nations, 2009) |
| | Closed broadleaved deciduous forest | *Class50* | |
| | Open broadleaved deciduous forest | *Class60* | |
| | Closed needleleaved evergreen forest | *Class70* | |
| | Open needleleaved deciduous or evergreen forest | *Class90* | |
| | Mixed broadleaved and needleleaved forest | *Class100* | |
| | Mosaic Forest/Shrubland / Grassland | *Class110* | |
| | Mosaic Grassland / Forest/Shrubland | *Class120* | |
| | Shrubland | *Class130* | |
| | Grassland | *Class140* | |
| | Sparse vegetation | *Class150* | |
| | Broadleaved forest regularly flooded - Fresh or brackish water | *Class160* | |
| | Broadleaved forest or shrubland permanently flooded - Saline or brackish water | *Class170* | |

| | | | |
|---|---|---|---|
| | Grassland or woody vegetation on regularly flooded or waterlogged soil - Fresh, brackish or saline water | *Class180* | |
| | Bare areas | *Class200* | |
| | Permanent snow and ice | *Class220* | |
| Hydrography | Distance to rivers | *D_riv* | Center for Global Environmental Research (2007) |
| Topography | Elevation | *Elevation* | US Geological Survey (1996) |
| | Slope | *Slope* | US Geological Survey (1996) |
| Spatial predictors | Spatial component where *x* refers to the biogeographical region stablish for modelling deer and tick species. | F*x* | Linear combination of latitude and longitude variables derived from trend surface analyses (Legendre, 1993) |
| Carrier predictors | Chorotype species richness, where *x* refers to the number of the chorotype. | SRx | Figure S2 |
| Amplifier predictors | Deer species models, where *x* refers to the species name. | M_*x* | Table S10 and Figure S5 |

**Table S3. Literature used to georeference the distribution of reports of Lyme borreliosis in humans**. Asterisks indicate countries for which the epidemiological information was provided by the Global Infectious Diseases and Epidemiology Network (GIDEON).

| COUNTRY | SOURCES |
|---|---|
| Albania | (Teita M. et al., 2018) |
| Algeria | (Bouattour et al., 2004) |
| Armenia | (Avagyang. A. et al., 2012) |
| Austria* | (Markowicz et al., 2021; Stanek et al., 1987; Stiasny et al., 2021) |
| Canada | (Gasmi et al., 2022; Henry et al., 2011; Public Health Agency of Canada, 2022; Tutt-Guérette et al., 2021) |
| China | (Che et al., 2022) |
| Belarus | (Leonidovna Sovkich et al., 2017) |
| Belgium* | (Bleyenheuft et al., 2015; Geebelen et al., 2019, 2022) |
| Bosnia and Herzegovina* | (Arapovic et al., 2014; Banović et al., 2021) |
| Bulgaria* | (Ermenlieva et al., 2019) |
| Croatia* | (Borčić et al., 1999; Topolovec et al., 2003) |
| Czech Republic* | (Kříž et al., 2018) |
| Denmark* | (Tetens et al., 2020) |
| Egypt* | (Elhelw et al., 2014) |
| Estonia* | (Tiina Prükk, 1999) |
| Finland* | (Kuitunen & Renko, 2022) |
| France* | (Petitgas et al., 2022) |
| Georgia | (Vashakidze et al., 2018) |
| Germany* | (Enkelmann et al., 2018; Hassenstein et al., 2022; Woudenberg et al., 2020) |
| Hungary* | (Zöldi et al., 2013; Zsuzsanna et al., 2019) |
| Ireland* | (Forde et al., 2021) |
| India* | (Babu et al., 2020; Vinayaraj et al., 2021) |
| Italy* | (Trevisan et al., 2023) |
| Japan* | (Masuzawa, 2004; Yamaji et al., 2018) |
| Kazakhstan* | (Head et al., 2020; Perfilyeva et al., 2020) |
| Latvia | (Kovalchuka et al., 2016) |
| Lithuania* | (Motiejunas et al., 1994) |
| Luxembourg* | (Reiffers-Mettelock et al., 1986) |

| | |
|---|---|
| Mexico* | (Colunga-Salas et al., 2020) |
| Moldova | (Caterinciuc et al., 2016) |
| Mongolia* | (Černý et al., 2019) |
| Montenegro | (Andrić et al., 2012) |
| Morocco | (Bouattour et al., 2004) |
| Nepal* | (Pun et al., 2018) |
| Netherlands* | (Coumou et al., 2011; Nassar-Sheikh Rashid et al., 2018) |
| North Macedonia | (Hadži-Petruševa Meloska & Hadži-Petruševa Jankijevič, 2014; Karageorgou et al., 2022) |
| Norway* | (Haugeberg et al., 2014; Nygård et al., 2005) |
| Poland* | (Brzozowska et al., 2021) |
| Portugal* | (Lopes de Carvalho & Núncio, 2006) |
| Romania* | (Kalmár et al., 2021) |
| Russian Federation | (Magnaval et al., 2016; Rudakova et al., 2021) |
| Serbia* | (Antonijević et al., 1993; Djerković et al., 1993; Stefanović et al., 1993) |
| Slovakia* | (Bušová et al., 2018; Svihrova et al., 2011) |
| Slovenia* | (Strle, 1999) |
| South Korea* | (Im et al., 2018; Moon et al., 2013; Seo, 2019) |
| Spain* | (Barreiro-Hurlé et al., 2020; Oteiza-Olaso et al., 2011; Vázquez-López et al., 2015) |
| Sweden* | (Wilhelmsson et al., 2016) |
| Switzerland* | (Altpeter et al., 2013) |
| Turkey* | (Önal et al., 2019) |
| Tunisia* | (Perveen et al., 2021; Younsi et al., 2001) |
| Ukraine* | (Rogovskyy et al., 2020) |
| United Kingdom | (Dubrey et al., 2014; Mavin et al., 2009; Tulloch et al., 2019) |

| | |
|---|---|
| United States of America | Literature from GIDEON:<br>(Avner, 2023; Bisanzio et al., 2020; Brown Marusiak et al., 2022; Brummitt et al., 2020; Clark & Villegas Nunez, 2023; Doggett et al., 2008; Dykstra et al., 2020; Maxwell et al., 2022; Pasternak A. et al., 2021; Stone et al., 2015; Witmer et al., 2022)<br><br>Epidemiological information from Departments of Public Health:<br>(Georgia Department of Public Health, 2023; Illinois Department of Public Health, 2023; Indiana Department of Health, 2023; Iowa Public Health Tracking Portal, 2021; Oklahoma State Department of Health, 2023; South Carolina Department of Health and Environmental Control, 2023; Tennessee Department of Health, 2021)<br><br>Other sources:<br>(CDC, 2022d; ProMED-mail, 2016a, 2017c, 2021b) |

**Table S4. Literature used to georeference the introduced free-living population of deer in Europe and North America.**

| DEER SPECIES WITH INTRODUCED POPULATION | SOURCES |
|---|---|
| *Cervus nippon* | (Feldhamer & Demarais, 2009; Ludek Bartos, 2009; Mattioli, 2011) |
| *Dama dama* | (Feldhamer & Demarais, 2009) |
| *Hydropotes inermis* | (Mattioli, 2011; Putman et al., 2021; Schilling & Rössner, 2017) |
| *Muntiacus reevesi* | (Baiwy et al., 2013; Mattioli, 2011) |
| *Odocoileus virginianus* | (Mattioli, 2011) |

**Table S5. Vector species models' logit functions (i.e., linear combinations of predictor variables in the probability equations obtained with logistic regression).** B: variable coefficient; SE: standard error; W: Wald parameter; DF: degrees of freedom; S: statistical significance. $X^2$ values and significances are provided for every model. Variable codes as in Table S2.

| *Ixodes ricinus* | | | | | |
|---|---|---|---|---|---|
| **Variable** | **B** | **SE** | **W** | **DF** | **S** |
| Bio15 | -0.079 | 0.016 | 24.253 | 1 | $8.447 \times 10^{-7}$ |
| Class120 | 19.870 | 7.036 | 7.975 | 1 | 0.005 |
| Class200 | 5.170 | 1.561 | 10.961 | 1 | $9.303 \times 10^{-4}$ |
| Pop_den | $-4.587 \times 10^{-5}$ | $1.463 \times 10^{-5}$ | 9.825 | 1 | 0.002 |
| Constant | 2.761 | 0.659 | 17.577 | 1 | $2.759 \times 10^{-5}$ |
| **Model goodness of fit** | $X^2 = 100.646$; $p < 0.05$ | | | | |
| *Ixodes persulcatus* | | | | | |
| **Variable** | **B** | **SE** | **W** | **DF** | **S** |
| Bio6 | -0.022 | 0.003 | 43.149 | 1 | $5.073 \times 10^{-11}$ |
| Class100 | 13.745 | 3.119 | 19.426 | 1 | $1.046 \times 10^{-5}$ |
| Bio5 | 0.013 | 0.005 | 7.738 | 1 | 0.005 |
| Constant | -7.491 | 1.570 | 22.778 | 1 | $1.818 \times 10^{-6}$ |
| **Model goodness of fit** | $X^2 = 113.603$; $p < 0.05$ | | | | |
| *Ixodes scapularis* | | | | | |
| **Variable** | **B** | **SE** | **W** | **DF** | **S** |
| Slope | -1.668 | 0.520 | 10.305 | 1 | 0.001 |
| Class50 | 6.777 | 2.230 | 9.239 | 1 | 0.002 |
| Constant | 1.566 | 0.707 | 4.911 | 1 | 0.027 |
| **Model goodness of fit** | $X^2 = 60.132$; $p < 0.05$ | | | | |
| *Ixodes pacificus* | | | | | |
| **Variable** | **B** | **SE** | **W** | **DF** | **S** |
| Slope | 1.189 | 0.485 | 6.021 | 1 | 0.014 |
| Class130 | 5.727 | 2.414 | 5.631 | 1 | 0.018 |
| Class90 | 15.076 | 5.535 | 7.420 | 1 | 0.006 |
| Constant | -6.272 | 1.837 | 11.662 | 1 | $6.380 \times 10^{-4}$ |
| **Model goodness of fit** | $X^2 = 27.706$; $p < 0.05$ | | | | |
| *Ixodes angustus* | | | | | |
| **Nearctic region** | | | | | |
| **Variable** | **B** | **SE** | **W** | **DF** | **S** |
| Class20 | -27.193 | 13.201 | 4.243 | 1 | 0.039 |
| F*Neartic* | 6.110 | 1.315 | 21.588 | 1 | $3.000 \times 10^{-6}$ |
| Constant | -2.319 | 0.765 | 9.204 | 1 | 0.002 |
| **Model goodness of fit** | $X^2 = 54.792$; $p < 0.05$ | | | | |
| **Palearctic region** | | | | | |
| **Variable** | **B** | **SE** | **W** | **DF** | **S** |
| Class70 | 11.399 | 4.886 | 5.443 | 1 | 0.020 |
| Class90 | 8.037 | 2.957 | 7.386 | 1 | 0.007 |
| Constant | -6.828 | 1.629 | 17.573 | 1 | $2.800 \times 10^{-5}$ |
| **Model goodness of fit** | $X^2 = 10.507$; $p < 0.05$ | | | | |

| *Ixodes sinensis* | | | | | |
|---|---|---|---|---|---|
| **Variable** | **B** | **SE** | **W** | **DF** | **S** |
| Class60 | 201.136 | 165.334 | 1.480 | 1 | 0.224 |
| F*Paleartic* | 12.700 | 6.535 | 3.777 | 1 | 0.052 |
| Constant | -12.217 | 6.150 | 3.945 | 1 | 0.047 |
| **Model goodness of fit** | $X^2 = 50.955$; $p < 0.05$ | | | | |
| ***Ixodes uriae*** | | | | | |
| **Nearctic region** | | | | | |
| **Variable** | **B** | **SE** | **W** | **DF** | **S** |
| Class150 | 6.748 | 2.867 | 5.541 | 1 | 0.019 |
| Constant | -2.277 | .416 | 29.924 | 1 | $4.494 \times 10^{-8}$ |
| **Model goodness of fit** | $X^2 = 7.379$; $p < 0.05$ | | | | |
| **Palearctic region** | | | | | |
| **Variable** | **B** | **SE** | **W** | **DF** | **S** |
| Class220 | 8.375 | 6.060 | 1.910 | 1 | 0.167 |
| Bio15 | -0.079 | 0.024 | 10.684 | 1 | $1.081 \times 10^{-3}$ |
| Class20 | -12.914 | 4.042 | 10.210 | 1 | $1.397 \times 10^{-3}$ |
| Constant | 1.667 | 0.954 | 3.051 | 1 | $8.067 \times 10^{-2}$ |
| **Model goodness of fit** | $X^2 = 44.259$; $p < 0.05$ | | | | |
| ***Ixodes spinipalpis*** | | | | | |
| **Variable** | **B** | **SE** | **W** | **DF** | **S** |
| Elevation | 0.002 | 0.001 | 8.847 | 1 | 0.003 |
| Slope | 0.829 | 0.352 | 5.558 | 1 | 0.018 |
| Constant | -5.242 | 1.245 | 17.739 | 1 | $2.500 \times 10^{-5}$ |
| **Model goodness of fit** | $X^2 = 29.290$; $p < 0.05$ | | | | |
| ***Amblyomma americanum*** | | | | | |
| **Variable** | **B** | **SE** | **W** | **DF** | **S** |
| Horses | 0.011 | 0.004 | 7.006 | 1 | 0.008 |
| Slope | -1.358 | 0.356 | 14.560 | 1 | $1.358 \times 10^{-4}$ |
| Class70 | 8.534 | 4.108 | 4.315 | 1 | 0.038 |
| Constant | 0.064 | 0.637 | 0.010 | 1 | 0.920 |
| **Model goodness of fit** | $X^2 = 39.413$; $p < 0.05$ | | | | |
| ***Haemaphysalis concinna*** | | | | | |
| **Variable** | **B** | **SE** | **W** | **DF** | **S** |
| Bio7 | 0.014 | 0.002 | 34.695 | 1 | $3.856 \times 10^{-9}$ |
| Class14 | 2.551 | 1.133 | 5.070 | 1 | 0.024 |
| Class30 | 11.331 | 2.966 | 14.594 | 1 | $1.333 \times 10^{-4}$ |
| Constant | -6.489 | 0.964 | 45.302 | 1 | $1.689 \times 10^{-11}$ |
| **Model goodness of fit** | $X^2 = 62.726$; $p < 0.05$ | | | | |
| ***Haemaphysalis longicornis*** | | | | | |
| **Nearctic region** | | | | | |
| **Variable** | **B** | **SE** | **W** | **DF** | **S** |
| Bio15 | -0.138 | 0.044 | 9.877 | 1 | 0.002 |
| Constant | 2.175 | 0.861 | 6.383 | 1 | 0.012 |
| **Model goodness of fit** | $X^2 = 30.243$; $p < 0.05$ | | | | |

| Palearctic region | | | | | |
|---|---|---|---|---|---|
| **Variable** | **B** | **SE** | **W** | **DF** | **S** |
| Pigs | $3.654 \times 10^{-4}$ | $9.283 \times 10^{-5}$ | 15.491 | 1 | $8.291 \times 10^{-5}$ |
| Goats | $7.372 \times 10^{-4}$ | $3.288 \times 10^{-4}$ | 5.026 | 1 | 0.025 |
| Bio12 | 0.002 | $7.236 \times 10^{-4}$ | 8.655 | 1 | 0.003 |
| Bio15 | 0.018 | 0.008 | 5.962 | 1 | 0.015 |
| Class20 | -14.482 | 5.853 | 6.122 | 1 | 0.013 |
| Class40 | -8.131 | 4.725 | 2.961 | 1 | 0.085 |
| Elevation | $5.833 \times 10^{-4}$ | $2.225 \times 10^{-4}$ | 6.871 | 1 | 0.009 |
| Constant | -5.248 | 1.117 | 22.056 | 1 | $2.648 \times 10^{-6}$ |
| **Model goodness of fit** | $X^2 = 111.203$; $p < 0.05$ | | | | |
| ***Dermacentor andersoni*** | | | | | |
| **Variable** | **B** | **SE** | **W** | **DF** | **S** |
| Elevation | 0.003 | $1.499 \times 10^{-3}$ | 4.659 | 1 | 0.031 |
| Class130 | 23.908 | 11.952 | 4.002 | 1 | 0.045 |
| Bio15 | 0.058 | 0.033 | 3.120 | 1 | 0.077 |
| Constant | -6.899 | 2.308 | 8.940 | 1 | 0.003 |
| **Model goodness of fit** | $X^2 = 55.499$; $p < 0.05$ | | | | |
| ***Dermacentor occidentalis*** | | | | | |
| **Variable** | **B** | **SE** | **W** | **DF** | **S** |
| Goats | 0.018 | 0.011 | 2.847 | 1 | 0.092 |
| Constant | -3.958 | 0.816 | 23.550 | 1 | $1.000 \times 10^{-6}$ |
| **Model goodness of fit** | $X^2 = 6.217$; $p < 0.05$ | | | | |
| ***Dermacentor variabilis*** | | | | | |
| **Variable** | **B** | **SE** | **W** | **DF** | **S** |
| Class50 | 112.316 | 43.997 | 6.517 | 1 | 0.011 |
| Slope | -0.999 | 0.459 | 4.742 | 1 | 0.029 |
| Constant | 0.147 | 0.908 | 0.026 | 1 | 0.871 |
| **Model goodness of fit** | $X^2 = 61.050$; $p < 0.05$ | | | | |
| ***Dermacentor silvarum*** | | | | | |
| **Variable** | **B** | **SE** | **W** | **DF** | **S** |
| Bio7 | 0.024 | 0.004 | 35.078 | 1 | $3.167 \times 10^{-9}$ |
| Bio15 | 0.019 | 0.005 | 13.546 | 1 | $2.328 \times 10^{-4}$ |
| Constant | -12.062 | 1.782 | 45.797 | 1 | $1.312 \times 10^{-11}$ |
| **Model goodness of fit** | $X^2 = 81.622$; $p < 0.05$ | | | | |
| ***Dermacentor nuttalli*** | | | | | |
| **Variable** | **B** | **SE** | **W** | **DF** | **S** |
| Bio7 | 0.020 | 0.004 | 23.771 | 1 | $1.085 \times 10^{-6}$ |
| Bio15 | 0.024 | 0.006 | 13.191 | 1 | $2.813 \times 10^{-4}$ |
| Elevation | 0.001 | 0.000 | 11.365 | 1 | $7.484 \times 10^{-4}$ |
| Constant | -12.226 | 2.040 | 35.910 | 1 | $2.067 \times 10^{-9}$ |
| **Model goodness of fit** | $X^2 = 61.067$; $p < 0.05$ | | | | |

**Table S6. Disease model's logit functions (i.e., linear combinations of predictor variables in the probability equations obtained with logistic-regression).** Disease models (DM) were made based on different predictors: environment, DM(e); carriers, DM(c); both environment and carriers, DM(e,c); both environment and vectors, DM(e,VM); and environment, carriers and vectors, DM(e,c,VM). B: variable coefficient; SE: standard error; W: Wald parameter; DF: degrees of freedom; S: statistical significance. $X^2$ values and significances are provided for every model. Variable codes as in Table S2. Species composition of chorotypes in the Figure S2.

| **DM (e)** | | | | | |
|---|---|---|---|---|---|
| **Variables** | **B** | **SE** | **W** | **DF** | **S** |
| Bio6 | -0.009 | 0.002 | 15.890 | 1 | $6.74x10^{-5}$ |
| Dis_rail | $-5.35x10^{-6}$ | $1.90x10^{-6}$ | 7.913 | 1 | 0.005 |
| Slope | -0.241 | 0.091 | 6.951 | 1 | 0.008 |
| Class20 | 4.692 | 1.714 | 7.491 | 1 | 0.006 |
| Class50 | 4.148 | 1.241 | 11.171 | 1 | 0.001 |
| Class70 | 7.526 | 2.438 | 9.527 | 1 | 0.002 |
| M_*Ccapygargus* | 2.689 | 0.808 | 11.078 | 1 | 0.001 |
| M_*Ddama* | 3.165 | 0.906 | 12.195 | 1 | $4.59x10^{-4}$ |
| M_*Mreevesi* | 2.027 | 0.798 | 6.458 | 1 | 0.011 |
| Constant | -1.153 | 0.437 | 6.963 | 1 | 0.008 |
| **Model goodness of fit** | $X^2$ = 200.780; $p < 0.05$ | | | | |
| **DM (c)** | | | | | |
| **Variables** | **B** | **SE** | **W** | **DF** | **S** |
| SR19 | 0.530 | 0.239 | 4.925 | 1 | 0.026 |
| SR30 | 0.175 | 0.029 | 37.440 | 1 | $9.43x10^{-10}$ |
| SR48 | 0.463 | 0.122 | 14.480 | 1 | $1.42x10^{-4}$ |
| SR56 | 0.194 | 0.032 | 35.700 | 1 | $2.30x10^{-9}$ |
| Constant | -1.574 | 0.255 | 38.083 | 1 | $6.78x10^{-10}$ |
| **Model goodness of fit** | $X^2$ = 155.028; $p < 0.05$ | | | | |
| **DM (e,c)** | | | | | |
| **Variables** | **B** | **SE** | **W** | **DF** | **S** |
| Bio6 | -0.013 | 0.003 | 15.186 | 1 | $9.786x10^{-5}$ |
| Dis_rail | $-7.884x10^{-6}$ | $2.414x10^{-6}$ | 10.667 | 1 | 0.001 |
| Slope | -0.073 | 0.107 | 0.456 | 1 | 0.499 |
| Class20 | -1.946 | 2.563 | 0.577 | 1 | 0.448 |
| Class50 | 1.942 | 1.475 | 1.732 | 1 | 0.188 |
| Class70 | 11.323 | 3.008 | 14.175 | 1 | $1.673x10^{-4}$ |
| *M_Ccapygargus* | 3.357 | 0.911 | 13.590 | 1 | $2.274x10^{-4}$ |
| M_*Ddama* | 2.883 | 1.349 | 4.568 | 1 | 0.033 |
| M_*Mreevesi* | 3.202 | 0.923 | 12.037 | 1 | 0.001 |
| SR19 | 0.761 | 0.349 | 4.755 | 1 | 0.029 |
| SR30 | 0.151 | 0.059 | 6.569 | 1 | 0.010 |
| SR44 | 1.225 | 0.307 | 15.896 | 1 | $6.709x10^{-5}$ |
| SR56 | 0.126 | 0.051 | 6.093 | 1 | 0.014 |
| Constant | -3.129 | 0.740 | 17.871 | 1 | $2.379x10^{-5}$ |
| **Model goodness of fit** | $X^2$ = 233.394; $p < 0.05$ | | | | |
| **DM (e,*Ixodes*-VM)** | | | | | |
| **Variables** | **B** | **SE** | **W** | **DF** | **S** |
| Bio7 | 0.006 | 0.003 | 3.465 | 1 | 0.063 |

| | | | | | |
|---|---|---|---|---|---|
| Dis_rail | $-8.036 \times 10^{-6}$ | $1.874 \times 10^{-6}$ | 18.385 | 1 | $1.805 \times 10^{-5}$ |
| Class70 | 8.420 | 2.535 | 11.029 | 1 | $8.971 \times 10^{-4}$ |
| Class130 | -8.112 | 2.537 | 10.226 | 1 | $1.385 \times 10^{-3}$ |
| M_*Aalces* | 1.215 | 0.531 | 5.222 | 1 | 0.022 |
| M_*Celaphus* | 2.598 | 0.648 | 16.052 | 1 | $6.164 \times 10^{-5}$ |
| M_*Ccapygargus* | 2.523 | 0.881 | 8.196 | 1 | 0.004 |
| M_*Ixodes*-VM | 2.054 | 0.707 | 8.427 | 1 | 0.004 |
| Constant | -3.093 | 1.028 | 9.054 | 1 | 0.003 |
| **Model goodness of fit** | $X^2 = 200.153$; $p < 0.05$ | | | | |
| **DM (e,Tick-VM)** | | | | | |
| **Variables** | **B** | **SE** | **W** | **DF** | **S** |
| Bio7 | 0.009 | 0.003 | 8.476 | 1 | 0.004 |
| Dis_rail | $-5.491 \times 10^{-6}$ | $1.864 \times 10^{-6}$ | 8.680 | 1 | 0.003 |
| Class20 | 4.794 | 1.777 | 7.273 | 1 | 0.007 |
| Class70 | 6.548 | 2.642 | 6.144 | 1 | 0.013 |
| Class100 | 10.596 | 3.695 | 8.224 | 1 | 0.004 |
| Class130 | -6.695 | 2.458 | 7.419 | 1 | 0.006 |
| *M_Ccapygargus* | 1.808 | 0.863 | 4.390 | 1 | 0.036 |
| M_*Ddama* | 3.314 | 0.999 | 10.994 | 1 | $9.139 \times 10^{-4}$ |
| M_Tick-VM | 2.603 | 0.929 | 7.856 | 1 | 0.005 |
| *Constante* | -5.425 | 1.231 | 19.423 | 1 | $1.047 \times 10^{-5}$ |
| **Model goodness of fit** | $X^2 = 205.485$; $p < 0.05$ | | | | |
| **DM (e,c,*Ixodes*-VM)** | | | | | |
| **Variables** | **B** | **SE** | **W** | **DF** | **S** |
| Bio7 | 0.007 | 0.004 | 4.175 | 1 | 0.041 |
| Dis_rail | $-6.460 \times 10^{-6}$ | $2.135 \times 10^{-6}$ | 9.153 | 1 | 0.002 |
| Class70 | 12.432 | 3.055 | 16.562 | 1 | $4.709 \times 10^{-5}$ |
| Class130 | -5.717 | 2.603 | 4.822 | 1 | 0.028 |
| M_*Aalces* | 0.776 | 0.649 | 1.431 | 1 | 0.232 |
| M_*Celaphus* | 1.552 | 0.769 | 4.072 | 1 | 0.044 |
| M_*Ccapygargus* | 3.095 | 1.012 | 9.352 | 1 | 0.002 |
| *Ixodes*-VM | 2.541 | 0.955 | 7.082 | 1 | 0.008 |
| SR30 | 0.165 | 0.043 | 14.648 | 1 | $1.295 \times 10^{-4}$ |
| SR44 | 0.813 | 0.256 | 10.046 | 1 | 0.002 |
| SR56 | 0.080 | 0.039 | 4.108 | 1 | 0.043 |
| Constante | -5.799 | 1.496 | 15.020 | 1 | $1.064 \times 10^{-4}$ |
| **Model goodness of fit** | $X^2 = 224.484$; $p > 0.05$ | | | | |
| **DM (e,c,Tick-VM)** | | | | | |
| **Variables** | **B** | **SE** | **W** | **DF** | **S** |
| Bio7 | 0.009 | 0.004 | 6.201 | 1 | 0.013 |
| Dis_rail | $-6.797 \times 10^{-6}$ | $2.009 \times 10^{-6}$ | 11.448 | 1 | $7.159 \times 10^{-4}$ |
| Class20 | 0.953 | 2.575 | 0.137 | 1 | 0.711 |
| Class70 | 7.977 | 2.842 | 7.880 | 1 | 0.005 |
| Class100 | 9.649 | 4.447 | 4.708 | 1 | 0.030 |
| Class130 | -6.380 | 2.380 | 7.190 | 1 | 0.007 |
| M_*Ccapygargus* | 2.301 | 0.985 | 5.459 | 1 | 0.019 |
| M_*Ddama* | 5.090 | 1.335 | 14.542 | 1 | $1.371 \times 10^{-4}$ |

| | | | | | |
|---|---|---|---|---|---|
| Tick-VM | 5.270 | 1.330 | 15.701 | 1 | $7.420x10^{-5}$ |
| SR19 | 0.943 | 0.339 | 7.752 | 1 | 0.005 |
| SR44 | 1.288 | 0.319 | 16.267 | 1 | $5.501x10^{-5}$ |
| Constante | -7.978 | 1.783 | 20.021 | 1 | $7.661x10^{-6}$ |
| **Model goodness of fit** | $X^2 = 232.749$; $p > 0.05$ | | | | |

**Table S7. Assessment of tick and deer species models based on discrimination and classification capacities.** The favourability threshold for classification was 0.5. PO: population origin, native (N) or introduced (I) population; AUC: area under the receiver operator characteristic curve; FCT: favourability classification threshold; TSS: True Skill Statistic; Sens.: sensitivity; Spec.: specificity; CCR: correct classification rate; Underp.: underprediction rate; Overp.: overprediction rate.

| TICK SPECIES MODELS | | | | | | | |
|---|---|---|---|---|---|---|---|
| **Model** | **AUC** | **TSS** | **Sens.** | **Spec.** | **CCR** | **Underp.** | **Overp.** |
| *Amblyomma americanum* | 0.912 | 0.678 | 0.789 | 0.889 | 0.838 | 0.200 | 0.118 |
| *Ixodes angustus* | 0.984 | 0.840 | 0.878 | 0.962 | 0.950 | 0.019 | 0.217 |
| *Ixodes pacificus* | 0.962 | 0.806 | 0.900 | 0.906 | 0.905 | 0.017 | 0.400 |
| *Ixodes persulcatus* | 0.950 | 0.697 | 0.831 | 0.866 | 0.856 | 0.072 | 0.289 |
| *Ixodes ricinus* | 0.936 | 0.625 | 0.813 | 0.812 | 0.812 | 0.082 | 0.373 |
| *Ixodes scapularis* | 0.998 | 0.842 | 0.911 | 0.931 | 0.919 | 0.129 | 0.047 |
| *Ixodes sinensis* | 0.993 | 0.941 | 1.000 | 0.941 | 0.943 | 0.000 | 0.591 |
| *Ixodes spinipalpis* | 0.973 | 0.785 | 0.833 | 0.952 | 0.932 | 0.033 | 0.231 |
| *Ixodes uriae* | 0.956 | 0.669 | 0.731 | 0.939 | 0.921 | 0.026 | 0.472 |
| *Dermacentor andersoni* | 0.993 | 0.927 | 0.944 | 0.982 | 0.973 | 0.018 | 0.056 |
| *Dermacentor nuttalli* | 0.978 | 0.806 | 0.864 | 0.942 | 0.934 | 0.015 | 0.387 |
| *Dermacentor occidentalis* | 0.995 | 0.986 | 1.000 | 0.986 | 0.986 | 0.000 | 0.250 |
| *Dermacentor silvarum* | 0.989 | 0.875 | 0.886 | 0.990 | 0.974 | 0.020 | 0.061 |
| *Dermacentor variabilis* | 0.987 | 0.811 | 0.811 | 1.000 | 0.865 | 0.323 | 0.000 |
| *Haemaphysalis concinna* | 0.875 | 0.557 | 0.684 | 0.873 | 0.808 | 0.160 | 0.260 |
| *Haemaphysalis longicornis* | 0.984 | 0.797 | 0.833 | 0.964 | 0.941 | 0.036 | 0.167 |

| DEER SPECIES MODELS | | | | | | | | |
|---|---|---|---|---|---|---|---|---|
| **Model** | **PO** | **AUC** | **TSS** | **Sens.** | **Spec.** | **CCR** | **Underp.** | **Overp.** |
| *Alces alces* | N | 0.991 | 0.892 | 0.940 | 0.952 | 0.947 | 0.038 | 0.076 |
| *Capreolus capreolus* | N | 0.993 | 0.897 | 0.943 | 0.953 | 0.950 | 0.024 | 0.108 |
| *Capreolus pygargus* | N | 0.985 | 0.791 | 0.817 | 0.974 | 0.937 | 0.054 | 0.094 |
| *Cervus albirostris* | N | 0.998 | 0.982 | 1.000 | 0.982 | 0.983 | 0.000 | 0.444 |
| *Cervus canadensis* | N | 0.991 | 0.890 | 0.946 | 0.943 | 0.944 | 0.013 | 0.209 |
| *Cervus elaphus* | N | 0.973 | 0.848 | 0.951 | 0.897 | 0.908 | 0.014 | 0.301 |
| *Cervus hanglu* | N | 0.986 | 0.555 | 0.571 | 0.983 | 0.974 | 0.010 | 0.556 |
| *Cervus nippon* | N | 0.960 | 0.622 | 0.700 | 0.922 | 0.913 | 0.015 | 0.708 |
| | I | 0.836 | 0.498 | 0.821 | 0.676 | 0.690 | 0.026 | 0.795 |
| *Dama dama* | N | 0.989 | 0.864 | 0.891 | 0.973 | 0.960 | 0.020 | 0.146 |
| *Dama mesopotamica* | N | 0.998 | 0.978 | 1.000 | 0.978 | 0.978 | 0.000 | 0.714 |
| *Hydropotes inermis* | N | 0.996 | 0.658 | 0.667 | 0.991 | 0.983 | 0.009 | 0.333 |
| | I | 0.992 | 0.982 | 1.000 | 0.987 | 0.983 | 0.000 | 0.571 |
| *Muntiacus reevesi* | N | 0.999 | 0.931 | 0.938 | 0.993 | 0.990 | 0.003 | 0.118 |
| | I | 0.984 | 0.907 | 1.000 | 0.907 | 0.908 | 0.000 | 0.875 |
| *Odocoileus hemionus* | N | 0.985 | 0.938 | 0.958 | 0.980 | 0.973 | 0.020 | 0.042 |
| *Odocoileus virginianus* | N | 0.999 | 0.926 | 0.926 | 1.000 | 0.932 | 0.455 | 0.000 |
| | I | 0.981 | 0.969 | 1.000 | 0.969 | 0.969 | 0.000 | 0.700 |
| *Rangifer tarandus* | N | 0.997 | 0.920 | 0.944 | 0.976 | 0.970 | 0.012 | 0.105 |

**Table S8. Assessment of vector models (VM) and disease models (DM)**. The favourability threshold for classification was 0.5. AUC: area under the receiver operator characteristic curve; FCT: favourability classification threshold; TSS: True Skill Statistic; Sens.: sensitivity; Spec.: specificity; CCR: correct classification rate; Underp.: underprediction rate; Overp.: overprediction rate.

| **Model** | | **AUC** | **FTC** | **TSS** | **Sens.** | **Spec.** | **CCR** | **Underp.** | **Overp.** |
|---|---|---|---|---|---|---|---|---|---|
| Disease Model | DM(e) | 0.935 | 0.5 | 0.687 | 0.859 | 0.829 | 0.848 | 0.243 | 0.096 |
| | DM(c) | 0.891 | 0.5 | 0.680 | 0.823 | 0.857 | 0.835 | 0.280 | 0.084 |
| | DM(e,c) | 0.955 | 0.5 | 0.761 | 0.904 | 0.857 | 0.888 | 0.174 | 0.077 |
| | DM(e,*Ixodes*-VM) | 0.929 | 0.5 | 0.674 | 0.874 | 0.800 | 0.848 | 0.229 | 0.108 |
| | DM(e,Tick-VM) | 0.936 | 0.5 | 0.717 | 0.889 | 0.829 | 0.868 | 0.202 | 0.093 |
| Vector Model | *Ixodes*-VM | 0.852 | 0.5 | 0.483 | 0.946 | 0.538 | 0.785 | 0.135 | 0.240 |
| | Tick-VM | 0.902 | 0.5 | 0.488 | 0.971 | 0.516 | 0.832 | 0.111 | 0.181 |

**Table S9. Assessment of predictive capacity of the downscaled Lyme borreliosis transmission model.** The assessment was based on discrimination and classification capacities respect to georeferenced reports in China (Che et al., 2022) and USA (CDC, 2022d). The favourability threshold for classification was 0.5. AUC: area under the receiver operator characteristic curve; TSS: True Skill Statistic; Sens.: sensitivity; Spec.: specificity; CCR: correct classification rate; Underp.: underprediction rate; Overp.: overprediction rate.

| Country | AUC | TSS | Sens. | Spec. | CCR | Underp. | Overp. |
|---|---|---|---|---|---|---|---|
| China | 0.675 | 0.245 | 0.668 | 0.576 | 0.589 | 0.085 | 0.797 |
| USA | 0.764 | 0.473 | 0.797 | 0.675 | 0.709 | 0.103 | 0.515 |

**Table S10. Deer model's logit functions (i.e., linear combinations of predictor variables in the probability equations obtained with logistic regression).** The fragmented distribution of *Dama dama* in North America, due to its introduction by humans, has led us to consider the maximum favourability ($F$ = 1) in every spatial unit with presence in this continent. In the case of *Dama mesopotamica, Muntiacus reevesi* (outside its native distribution area), and *Cervus nippon* (outside its native distribution area in the Nearctic), only the spatial model was considered, as we found no significant environmental model. B: variable coefficient; SE: standard error; W: Wald parameter; DF: degrees of freedom; S: statistical significance. Variable codes as in Table S2.

| *Alces alces* | | | | | |
|---|---|---|---|---|---|
| **Variable** | **B** | **SE** | **W** | **DF** | **S** |
| Bio6 | -0.008 | 0.004 | 4.316 | 1 | 0.038 |
| Bio12 | -0.009 | 0.002 | 12.173 | 1 | $4.85 \times 10^{-04}$ |
| Class50 | 10.664 | 2.890 | 13.617 | 1 | $2.24 \times 10^{-04}$ |
| Class100 | 32.587 | 11.289 | 8.332 | 1 | 0.004 |
| Class120 | 18.586 | 7.920 | 5.507 | 1 | 0.019 |
| F*Nearctic* | 6.541 | 1.299 | 25.363 | 1 | $4.75 \times 10^{-07}$ |
| F*Palearctic* | 6.837 | 1.142 | 35.826 | 1 | $2.16 \times 10^{-09}$ |
| Constant | -2.738 | 0.991 | 7.631 | 1 | 0.006 |
| **Model goodness of fit** | $X^2$ = 330.836; $p < 0.05$ | | | | |
| *Cervus albirostris* | | | | | |
| **Variable** | **B** | **SE** | **W** | **DF** | **S** |
| Elevation | 0.003 | $1.12 \times 10^{-03}$ | 8.869 | 1 | 0.003 |
| Class60 | 165.012 | 123.671 | 1.780 | 1 | 0.182 |
| Class140 | -20.527 | 9.718 | 4.462 | 1 | 0.035 |
| Constant | -8.276 | 2.097 | 15.579 | 1 | $7.91 \times 10^{-05}$ |
| **Model goodness of fit** | $X^2$ =29.484; $p < 0.05$ | | | | |
| *Cervus canadensis* | | | | | |
| **Variable** | **B** | **SE** | **W** | **DF** | **S** |
| Class14 | -10.035 | 3.390 | 8.762 | 1 | 0.003 |
| F*Nearctic* | 9.706 | 1.635 | 35.247 | 1 | $2.90 \times 10^{-09}$ |
| F*Palearctic* | 8.657 | 1.474 | 34.488 | 1 | $4.29 \times 10^{-09}$ |
| Constant | -5.666 | 1.108 | 26.159 | 1 | $3.14 \times 10^{-07}$ |
| **Model goodness of fit** | $X^2$ =230.885; $p < 0.05$ | | | | |
| *Cervus elaphus* | | | | | |
| **Variable** | **B** | **SE** | **W** | **DF** | **S** |
| Bio15 | -0.046 | 0.011 | 18.662 | 1 | $1.60 \times 10^{-05}$ |
| Sheep | $5.59 \times 10^{-04}$ | $1.65 \times 10^{-04}$ | 11.488 | 1 | $7.00 \times 10^{-04}$ |
| P_den | $-6.30 \times 10^{-05}$ | $1.80 \times 10^{-05}$ | 12.098 | 1 | $5.05 \times 10^{-04}$ |
| Class20 | 3.415 | 1.223 | 7.793 | 1 | 0.005 |
| Class120 | 31.417 | 7.099 | 19.586 | 1 | $1.00 \times 10^{-05}$ |
| Constant | 0.182 | 0.603 | 0.091 | 1 | 0.763 |
| **Model goodness of fit** | $X^2$ =120.695; $p < 0.05$ | | | | |
| *Cervus hanglu* | | | | | |

| Variable | B | SE | W | DF | S |
|---|---|---|---|---|---|
| Elevation | 7.49x10$^{-04}$ | 2.49x10$^{-04}$ | 9.076 | 1 | 0.003 |
| Constant | -4.440 | 0.527 | 70.872 | 1 | 3.81x10$^{-17}$ |
| **Model goodness of fit** | $X^2$ =6.731; $p < 0.05$ | | | | |
| ***Capreolus capreolus*** | | | | | |
| **Variable** | **B** | **SE** | **W** | **DF** | **S** |
| Class110 | 8.292 | 3.963 | 4.378 | 1 | 0.036 |
| F*Palearctic* | 8.571 | 1.056 | 65.911 | 1 | 4.72x10$^{-16}$ |
| Constant | -5.393 | 0.837 | 41.481 | 1 | 1.19x10$^{-10}$ |
| **Model goodness of fit** | $X^2$ =293.157; $p < 0.05$ | | | | |
| ***Capreolus pygargus*** | | | | | |
| **Variable** | **B** | **SE** | **W** | **DF** | **S** |
| Bio7 | 0.031 | 0.004 | 57.199 | 1 | 3.94x10$^{-14}$ |
| Bio15 | 0.015 | 0.006 | 6.462 | 1 | 0.011 |
| Class14 | 8.996 | 1.658 | 29.448 | 1 | 5.75x10$^{-08}$ |
| Constant | -14.558 | 1.816 | 64.231 | 1 | 1.11x10$^{-15}$ |
| **Model goodness of fit** | $X^2$ =160.519; $p < 0.05$ | | | | |
| ***Odocoileus hemionus*** | | | | | |
| **Variable** | **B** | **SE** | **W** | **DF** | **S** |
| Class90 | 16.609 | 9.159 | 3.288 | 1 | 0.070 |
| F*Nearctic* | 10.346 | 3.331 | 9.648 | 1 | 0.002 |
| Constant | -7.166 | 2.941 | 5.938 | 1 | 0.015 |
| **Model goodness of fit** | $X^2$ =81.073; $p < 0.05$ | | | | |
| ***Rangifer tarandus*** | | | | | |
| **Nearctic region** | | | | | |
| **Variable** | **B** | **SE** | **W** | **DF** | **S** |
| Bio15 | -0.058 | 0.044 | 1.737 | 1 | 0.188 |
| P_den | 1.68x10$^{-04}$ | 1.16x10$^{-04}$ | 2.107 | 1 | 0.147 |
| Constant | 5.238 | 8.263 | 0.402 | 1 | 0.526 |
| **Model goodness of fit** | $X^2$ =60.935; $p < 0.05$ | | | | |
| **Palearctic region** | | | | | |
| **Variable** | **B** | **SE** | **W** | **DF** | **S** |
| Bio6 | -0.047 | 0.013 | 13.145 | 1 | 2.88x10$^{-04}$ |
| D_rail | 1.31x10$^{-05}$ | 4.43x10$^{-06}$ | 8.768 | 1 | 0.003 |
| Class20 | 12.350 | 4.281 | 8.322 | 1 | 0.004 |
| Class70 | 17.123 | 7.957 | 4.631 | 1 | 0.031 |
| Class90 | 10.734 | 3.971 | 7.305 | 1 | 0.007 |
| Class100 | 42.406 | 12.082 | 12.320 | 1 | 4.48x10$^{-04}$ |
| Class150 | 14.436 | 6.007 | 5.776 | 1 | 0.016 |
| Class180 | 28.982 | 13.121 | 4.879 | 1 | 0.027 |
| Class220 | 13.087 | 6.391 | 4.193 | 1 | 0.041 |
| Constant | -20.531 | 5.128 | 16.028 | 1 | 6.24x10$^{-05}$ |
| **Model goodness of fit** | $X^2$ =181.955; $p < 0.05$ | | | | |

| *Dama mesopotamica* | | | | | |
|---|---|---|---|---|---|
| **Variable** | **B** | **SE** | **W** | **DF** | **S** |
| La | 108.285 | 160.273 | 0.456 | 1 | 0.499 |
| Lo | 1.923 | 1.637 | 1.380 | 1 | 0.240 |
| $Lo^2$ | -0.017 | 0.015 | 1.349 | 1 | 0.245 |
| $La^3$ | -0.036 | 0.053 | 0.466 | 1 | 0.495 |
| Constant | -2323.115 | 3410.368 | 0.464 | 1 | 0.496 |
| **Model goodness of fit** | $X^2 = 17.739$; $p < 0.05$ | | | | |
| *Dama dama* | | | | | |
| **Native population distribution** | | | | | |
| **Variable** | **B** | **SE** | **W** | **DF** | **S** |
| Class120 | 49.358 | 10.790 | 20.925 | 1 | $4.78x10^{-06}$ |
| Bio7 | -0.006 | 0.003 | 3.605 | 1 | 0.058 |
| Bio15 | -0.046 | 0.013 | 13.651 | 1 | $2.20x10^{-04}$ |
| D_pop | $-1.77x10^{-4}$ | $3.89x10^{-05}$ | 20.597 | 1 | $5.67x10^{-06}$ |
| Class20 | 3.794 | 1.675 | 5.128 | 1 | 0.024 |
| Constant | 3.064 | 1.220 | 6.306 | 1 | 0.012 |
| **Model goodness of fit** | $X^2 =137.677$; $p < 0.05$ | | | | |
| *Muntiacus reevesi* | | | | | |
| **Native population distribution** | | | | | |
| **Variable** | **B** | **SE** | **W** | **DF** | **S** |
| Buffaloes | 0.003 | $1.12x10^{-03}$ | 5.508 | 1 | 0.019 |
| Pigs | $1.10x10^{-03}$ | $3.77x10^{-04}$ | 8.506 | 1 | 0.004 |
| AEI | -0.827 | 0.312 | 7.045 | 1 | 0.008 |
| Class11 | 37.648 | 15.160 | 6.168 | 1 | 0.013 |
| Class14 | 15.759 | 6.307 | 6.243 | 1 | 0.012 |
| Class50 | -204.638 | 86.717 | 5.569 | 1 | 0.018 |
| Class70 | 15.611 | 5.968 | 6.843 | 1 | 0.009 |
| Constant | -7.242 | 2.459 | 8.675 | 1 | 0.003 |
| **Model goodness of fit** | $X^2 =102.857$; $p < 0.05$ | | | | |
| **Introduced population distribution** | | | | | |
| **Variable** | **B** | **SE** | **W** | **DF** | **S** |
| $Lo^2$ | -0.026 | 0.016 | 2.520 | 1 | 0.112 |
| $La^3$ | $1,91x10^{-05}$ | $1,26x10^{-05}$ | 2.303 | 1 | 0.129 |
| Constant | -3.379 | 1.574 | 4.610 | 1 | 0.032 |
| **Model goodness of fit** | $X^2 =16.419$; $p < 0.05$ | | | | |
| *Odocoileus virginianus* | | | | | |
| **Native population distribution** | | | | | |
| **Variable** | **B** | **SE** | **W** | **DF** | **S** |
| Bio15 | 0.182 | 0.117 | 2.417 | 1 | 0.120 |
| D_riv | $-1.25x10^{-4}$ | $8.60x10^{-05}$ | 2.094 | 1 | 0.148 |
| Constant | -27.140 | 17.667 | 2.360 | 1 | 0.124 |
| **Model goodness of fit** | $X^2 =35.276$; $p < 0.05$ | | | | |

| Introduced population distribution | | | | | |
|---|---|---|---|---|---|
| **Variable** | **B** | **SE** | **W** | **DF** | **S** |
| Class120 | 16.080 | 8.972 | 3.212 | 1 | 0.073 |
| Constant | -5.034 | 0.849 | 35.186 | 1 | $3.00x10^{-09}$ |
| **Model goodness of fit** | $X^2$ =2.501; $p < 0.05$ | | | | |
| ***Hydropotes inermis*** | | | | | |
| **Native population distribution** | | | | | |
| **Variable** | **B** | **SE** | **W** | **DF** | **S** |
| Class11 | 8.954 | 2.256 | 15.760 | 1 | $7.20x10^{-05}$ |
| Bio12 | 0.002 | $7.71x10^{-04}$ | 3.961 | 1 | 0.047 |
| Constant | -5.915 | 1.148 | 26.566 | 1 | $2.55x10^{-07}$ |
| **Model goodness of fit** | $X^2$ =23.728; $p < 0.05$ | | | | |
| **Introduced population distribution** | | | | | |
| **Variable** | **B** | **SE** | **W** | **DF** | **S** |
| Sheep | $7.55x10^{-04}$ | $3.90x10^{-04}$ | 3.741 | 1 | 0.053 |
| Class120 | 32.460 | 15.703 | 4.273 | 1 | 0.039 |
| Class140 | 8.629 | 3.494 | 6.098 | 1 | 0.014 |
| Constant | -9.045 | 2.829 | 10.223 | 1 | $1.39x10^{-03}$ |
| **Model goodness of fit** | $X^2$ =18.940; $p < 0.05$ | | | | |
| ***Cervus nippon*** | | | | | |
| **Native population distribution** | | | | | |
| **Variable** | **B** | **SE** | **W** | **DF** | **S** |
| Class70 | 8.768 | 2.396 | 13.393 | 1 | $2.53x10^{-04}$ |
| Pigs | $1.65x10^{-04}$ | $6.09x10^{-05}$ | 7.360 | 1 | 0.007 |
| Constant | -4.361 | 0.590 | 54.561 | 1 | $1.51x10^{-13}$ |
| **Model goodness of fit** | $X^2$ = 18.591; $p < 0.05$ | | | | |
| **Introduced population distribution** | | | | | |
| **Nearctic region** | | | | | |
| **Variable** | **B** | **SE** | **W** | **DF** | **S** |
| Lo | 17.296 | 10.360 | 2.787 | 1 | 0.095 |
| $Lo^2$ | 0.175 | 0.109 | 2.584 | 1 | 0.108 |
| $La^2$ | -0.119 | 0.065 | 3.311 | 1 | 0.069 |
| LaxLo | -0.102 | 0.057 | 3.183 | 1 | 0.074 |
| $Lo^2xLa$ | $-1,10x10^{-03}$ | $6,15x10^{-04}$ | 3.192 | 1 | 0.074 |
| $La^2xLo$ | $-1,30x10^{-3}$ | $7,16x10^{-04}$ | 3.278 | 1 | 0.070 |
| $Lo^3$ | $5,13x10^{-04}$ | $3,49x10^{-04}$ | 2.156 | 1 | 0.142 |
| Constant | 505.003 | 299.852 | 2.836 | 1 | 0.092 |
| **Model goodness of fit** | $X^2$ =11.497; $p < 0.05$ | | | | |
| **Palearctic region** | | | | | |
| **Variable** | **B** | **SE** | **W** | **DF** | **S** |
| Bio15 | -0.070 | 0.022 | 9.775 | 1 | 0.002 |
| Constant | 0.336 | 0.714 | 0.222 | 1 | 0.638 |
| **Model goodness of fit** | $X^2$ =23.855; $p < 0.05$ | | | | |

**Figure S1. Tick vector distributions and favourability models. A.** Accumulated distribution of tick species belonging to genus *Ixodes*. **B.** Accumulated distribution of tick species belonging to genera *Ixodes*, *Dermacentor, Haemaphysalis*, and *Amblyomma*. **C.** *Ixodes* vector model resulting from the fuzzy union of favourability models of *Ixodes* species. **D.** Tick vector model resulting from the fuzzy union of favourability models of *Ixodes, Dermacentor, Haemaphysalis,* and *Amblyomma* species. Favourability was based on environmental and human predictors (see variables included in the models in Table S5). Favourability values were categorized as low (<0.2), low-intermediate (0.2-0.5), high-intermediate (0.5-0.8), and high (>0.8).

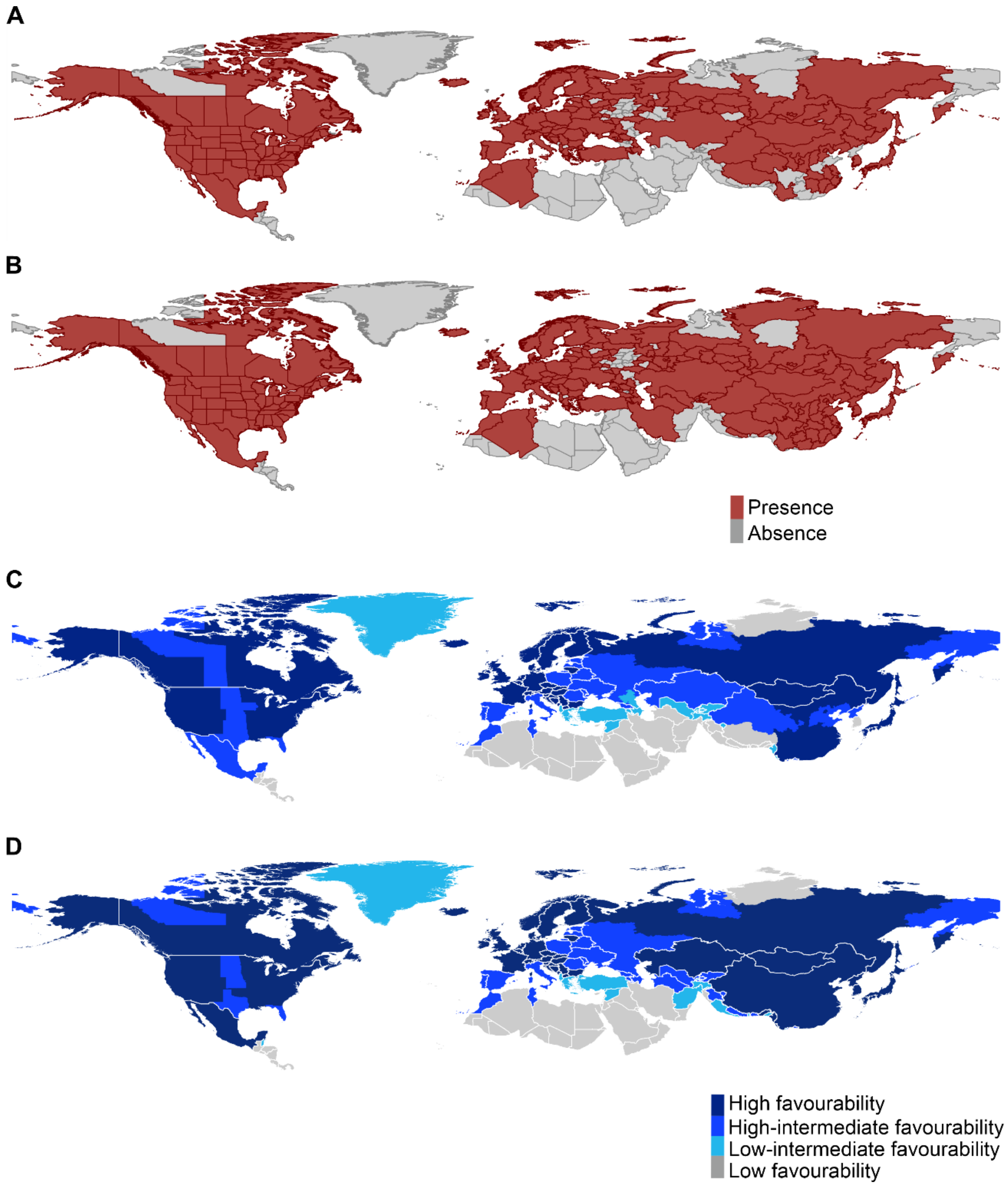

**Figure S2. Chorotypes considered for entry in the model after overcoming a false discovery rate test.** Maps represent the species richness of each chorotype projected into a hexagon grid (see downscaling section in the main text). The list of the species from each chorotype is also detailed. Chorotypes codes in bold have entered in both transmission models mention in the main text.

| CODE | CHOROTYPE | SPECIES |
|---|---|---|
| SR 05 | 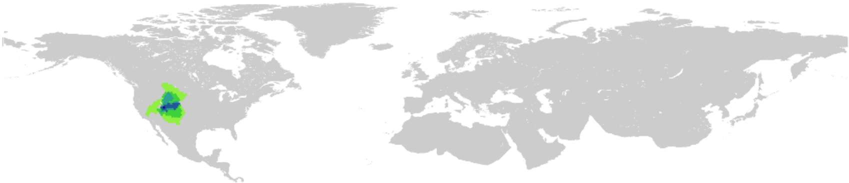 | *Cynomys leucurus*<br>*Cynomys parvidens*<br>*Neotamias quadrivittatus*<br>*Neotamias rufus*<br>*Neotamias umbrius*<br>*Sorex nanus* |
| SR 14 | 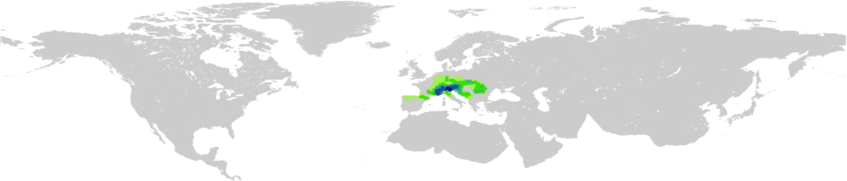 | *Apodemus alpicola*<br>*Arvicola scherman*<br>*Marmota marmota*<br>*Microtus liechtensteini*<br>*Microtus multiplex*<br>*Sorex alpinus* |
| **SR 19** | 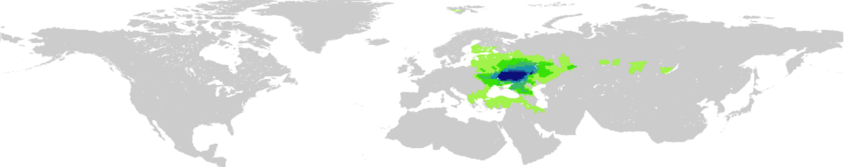 | *Desmana moschata*<br>*Microtus levis*<br>*Sicista severtzovi*<br>*Sicista strandi*<br>*Spalax microphthalmus*<br>*Spermophilus suslicus* |
| SR 22 | 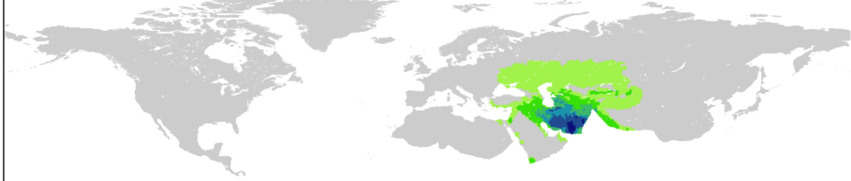 | *Allactaga major*<br>*Calomyscus hotsoni*<br>*Crocidura zarudnyi*<br>*Gerbillus aquilus*<br>*Hystrix indica*<br>*Jaculus blanfordi*<br>*Meriones persicus*<br>*Nesokia indica*<br>*Ochotona rufescens*<br>*Paraechinus hypomelas*<br>*Tatera indica* |
| SR 24 | 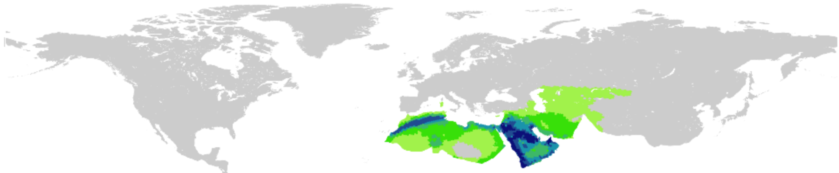 | *Acomys dimidiatus*<br>*Acomys russatus*<br>*Gerbillus cheesmani*<br>*Gerbillus dasyurus*<br>*Gerbillus henleyi*<br>*Gerbillus nanus*<br>*Jaculus jaculus*<br>*Lepus capensis*<br>*Meriones crassus*<br>*Meriones libycus*<br>*Paraechinus aethiopicus*<br>*Psammomys obesus* |
| **SR 30** | 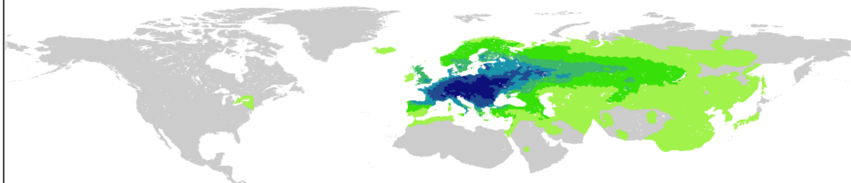 | *Apodemus agrarius*<br>*Apodemus flavicollis*<br>*Apodemus sylvaticus*<br>*Arvicola amphibius*<br>*Castor fiber*<br>*Crocidura leucodon*<br>*Crocidura suaveolens*<br>*Dryomys nitedula*<br>*Eliomys quercinus*<br>*Erinaceus europaeus*<br>*Erinaceus roumanicus*<br>*Glis glis*<br>*Lepus europaeus*<br>*Micromys minutus*<br>*Microtus agrestis*<br>*Microtus arvalis*<br>*Microtus subterraneus*<br>*Muscardinus avellanarius*<br>*Myodes glareolus*<br>*Neomys anomalus*<br>*Neomys fodiens*<br>*Oryctolagus cuniculus*<br>*Sicista betulina*<br>*Sorex araneus*<br>*Sorex minutus*<br>*Talpa europaea* |

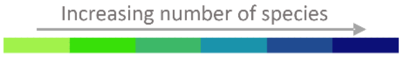

| CODE | CHOROTYPE | SPECIES |
| --- | --- | --- |
| SR 31 | 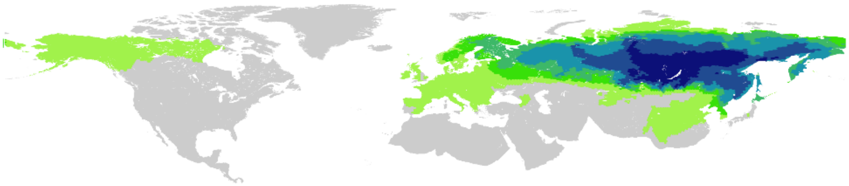 | *Apodemus peninsulae*<br>*Eutamias sibiricus*<br>*Lepus timidus*<br>*Microtus oeconomus*<br>*Myodes rufocanus*<br>*Myodes rutilus*<br>*Myopus schisticolor*<br>*Ochotona hyperborea*<br>*Ochotona turuchanensis*<br>*Pteromys volans*<br>*Sciurus vulgaris*<br>*Sorex caecutiens*<br>*Sorex daphaenodon*<br>*Sorex isodon*<br>*Sorex minutissimus*<br>*Sorex roboratus*<br>*Sorex tundrensis*<br>*Talpa altaica* |
| SR 37 | 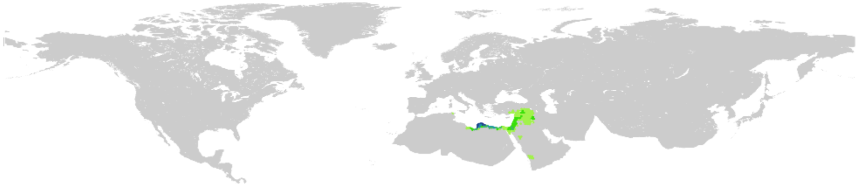 | *Allactaga tetradactyla*<br>*Crocidura aleksandrisi*<br>*Eliomys melanurus*<br>*Gerbillus andersoni*<br>*Gerbillus grobbeni*<br>*Nannospalax ehrenbergi* |
| SR 38 | 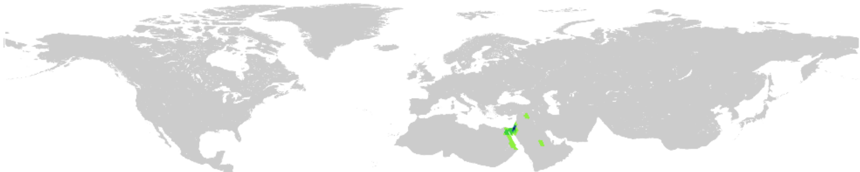 | *Crocidura katinka*<br>*Crocidura ramona*<br>*Gerbillus floweri*<br>*Meriones sacramenti*<br>*Sekeetamys calurus* |
| SR 41 | 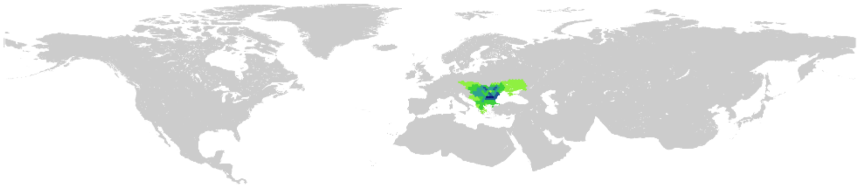 | *Mesocricetus newtoni*<br>*Microtus tatricus*<br>*Mus spicilegus*<br>*Nannospalax leucodon*<br>*Spalax graceus*<br>*Spermophilus citellus* |
| SR 42 | 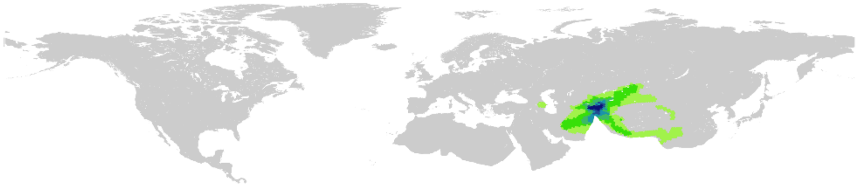 | *Alticola argentatus*<br>*Apodemus pallipes*<br>*Blanfordimys bucharensis*<br>*Crocidura gmelini*<br>*Crocidura serezkyensis*<br>*Ellobius alaicus*<br>*Marmota caudata*<br>*Neodon juldaschi*<br>*Ochotona rutila*<br>*Rattus pyctoris*<br>*Sorex buchariensis* |
| **SR 44** | 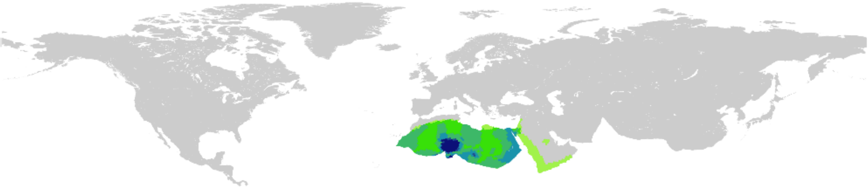 | Acomys airensis<br>*Acomys cahirinus*<br>*Acomys seurati*<br>*Gerbillus gerbillus*<br>*Gerbillus pyramidum*<br>*Gerbillus tarabuli*<br>*Massoutiera mzabi*<br>*Procavia capensis* |

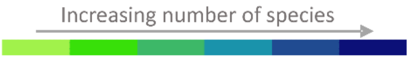

| CODE | CHOROTYPE | SPECIES |
|---|---|---|
| **SR 48** | 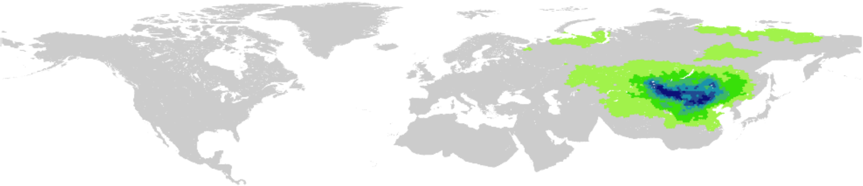 | *Allactaga balikunica*<br>*Allactaga bullata*<br>*Allocricetulus curtatus*<br>*Alticola barakshin*<br>*Alticola semicanus*<br>*Cardiocranius paradoxus*<br>*Cricetulus barabensis*<br>*Cricetulus longicaudatus*<br>*Cricetulus sokolovi*<br>*Crocidura sibirica*<br>*Eolagurus przewalskii*<br>*Lasiopodomys brandtii*<br>*Marmota sibirica*<br>*Meriones unguiculatus*<br>*Mesechinus dauuricus*<br>*Microtus gregalis*<br>*Microtus maximowiczii*<br>*Microtus mongolicus*<br>*Myospalax aspalax*<br>*Myospalax psilurus*<br>*Ochotona dauurica*<br>*Ochotona pallasii*<br>*Phodopus campbelli*<br>*Phodopus roborovskii*<br>*Salpingotus crassicauda*<br>*Spermophilus alashanicus*<br>*Spermophilus dauricus*<br>*Spermophilus pallidicauda*<br>*Stylodipus andrewsi*<br>*Urocitellus undulatus* |
| SR 54 | 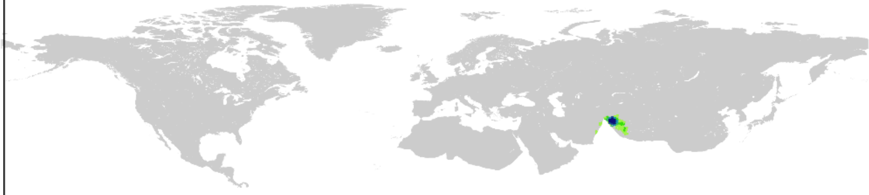 | *Alticola montosa*<br>*Apodemus rusiges*<br>*Crocidura pergrisea*<br>*Crocidura pullata*<br>*Eoglaucomys fimbriatus*<br>*Eupetaurus cinereus*<br>*Hyperacrius fertilis*<br>*Hyperacrius wynnei*<br>*Sorex planiceps* |
| SR 55 | 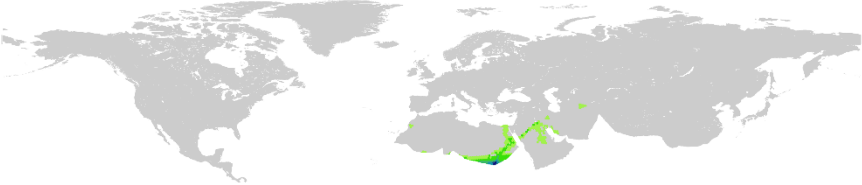 | *Acomys cineraceus*<br>*Allactaga euphratica*<br>*Crocidura fulvastra*<br>*Crocidura olivieri*<br>*Desmodilliscus braueri*<br>*Gerbilliscus robustus*<br>*Gerbillus muriculus*<br>*Gerbillus nancillus*<br>*Gerbillus principulus*<br>*Mastomys kollmannspergeri*<br>*Microtus guentheri*<br>*Microtus irani*<br>*Taterillus emini*<br>*Xerus erythropus* |
| **SR 56** | 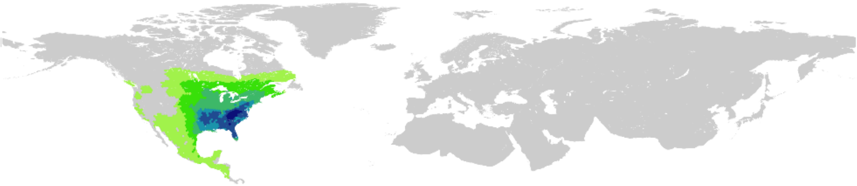 | *Blarina brevicauda*<br>*Blarina carolinensis*<br>*Blarina hylophaga*<br>*Condylura cristata*<br>*Cryptotis parva*<br>*Geomys breviceps*<br>*Geomys pinetis*<br>*Glaucomys volans*<br>*Microtus chrotorrhinus*<br>*Microtus pinetorum*<br>*Napaeozapus insignis*<br>*Neofiber alleni*<br>*Neotoma floridana*<br>*Neotoma magister*<br>*Ochrotomys nuttalli*<br>*Oryzomys palustris*<br>*Parascalops breweri*<br>*Peromyscus attwateri*<br>*Peromyscus aztecus*<br>*Peromyscus difficilis*<br>*Peromyscus gossypinus*<br>*Peromyscus leucopus*<br>*Peromyscus polionotus*<br>*Podomys floridanus*<br>*Reithrodontomys humulis*<br>*Scalopus aquaticus*<br>*Sciurus carolinensis*<br>*Sciurus niger*<br>*Sigmodon hispidus*<br>*Sorex dispar*<br>*Sorex fumeus*<br>*Sorex longirostris*<br>*Sylvilagus aquaticus*<br>*Sylvilagus floridanus*<br>*Sylvilagus obscurus*<br>*Sylvilagus palustris*<br>*Synaptomys cooperi*<br>*Tamias striatus* |

**Figure S3. Lyme borreliosis models.** Distribution of Lyme borreliosis in humans used as the dependent variable for disease model (DM) calibration. These models quantify if conditions are favourable to the presence of Lyme borreliosis in humans, based on different predictors: environment, **DM(e)**; carriers, **DM(c)**; environment and carriers, **DM(e,c)**; environment and favourability conditions for the presence of *Ixodes* tick species, **DM(e,*Ixodes*-VM)**; environment and favourability conditions for the presence of tick species, **DM(e,tick-VM)**; environment, carriers and favourability conditions for the presence of *Ixodes* species, **DM(e,c,*Ixodes*-VM)**; and, environment, carriers and favourability conditions for the presence of tick species, **DM(e,c,tick-VM)** (see variables included in the models in Table S2). Favourability values were categorized as low (<0.2), low-intermediate (0.2-0.5), high-intermediate (0.5-0.8), and high (>0.8).

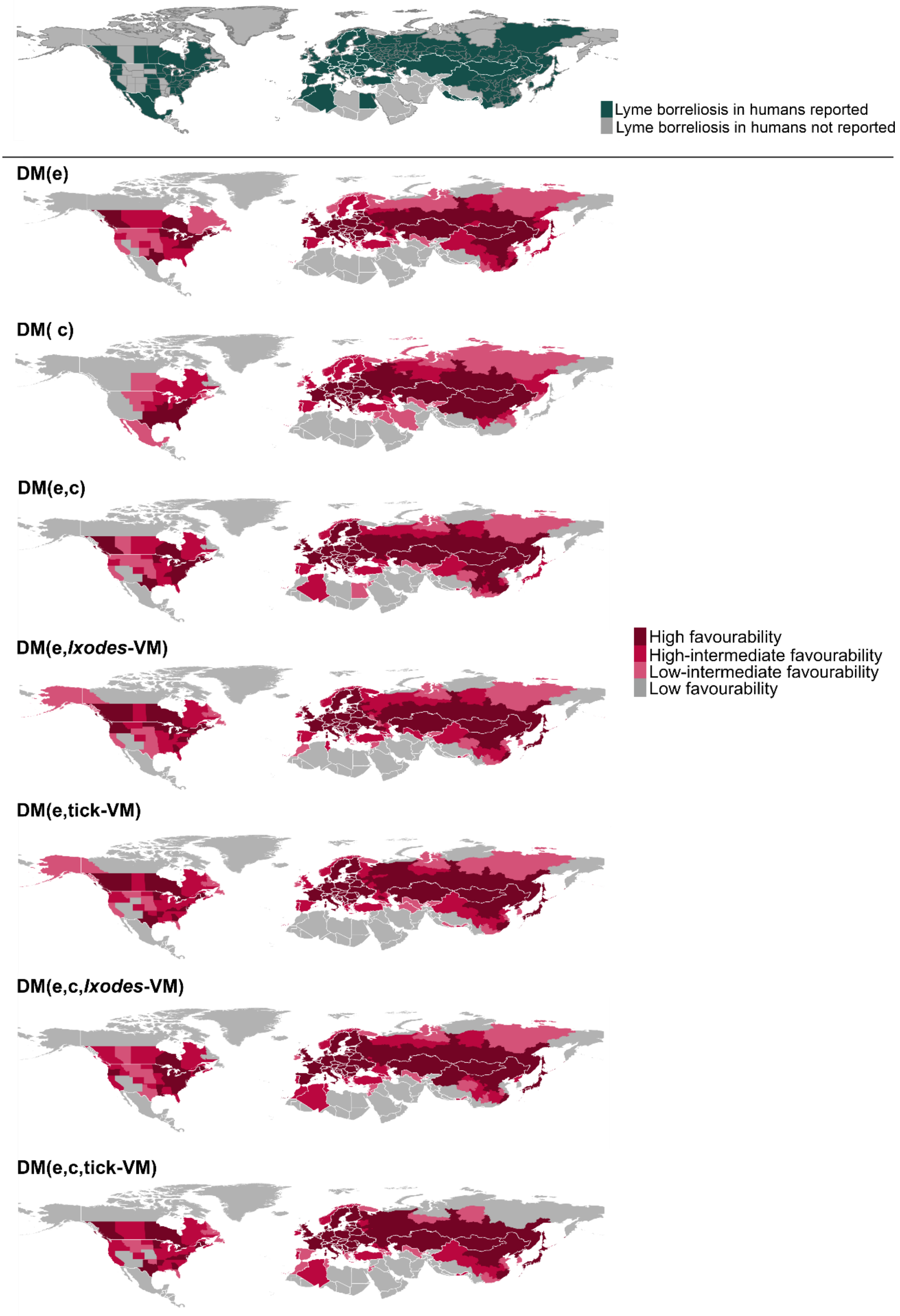

**Figure S4. Lyme borreliosis transmission risk models according to different pathogeographic scenarios. VCL scenario:** both vectors and wild carriers are geographically limiting factors. **CL scenario:** only carriers are limiting factors. **VL scenario:** only vectors are limiting factors. **NL scenario:** neither vectors nor carriers are limiting factors, i.e. vector and carriers are mutually compensating factors. The distribution of reports of Lyme borreliosis in humans is shown. *: Only *Ixodes* tick species are considered in the vector model; +: *Ixodes, Haemaphysalis, Dermacentor* and *Amblyomma* tick species are considered in vector model. Risk values were categorized as low (<0.2), low-intermediate (0.2-0.5), high-intermediate (0.5-0.8), and high (>0.8).

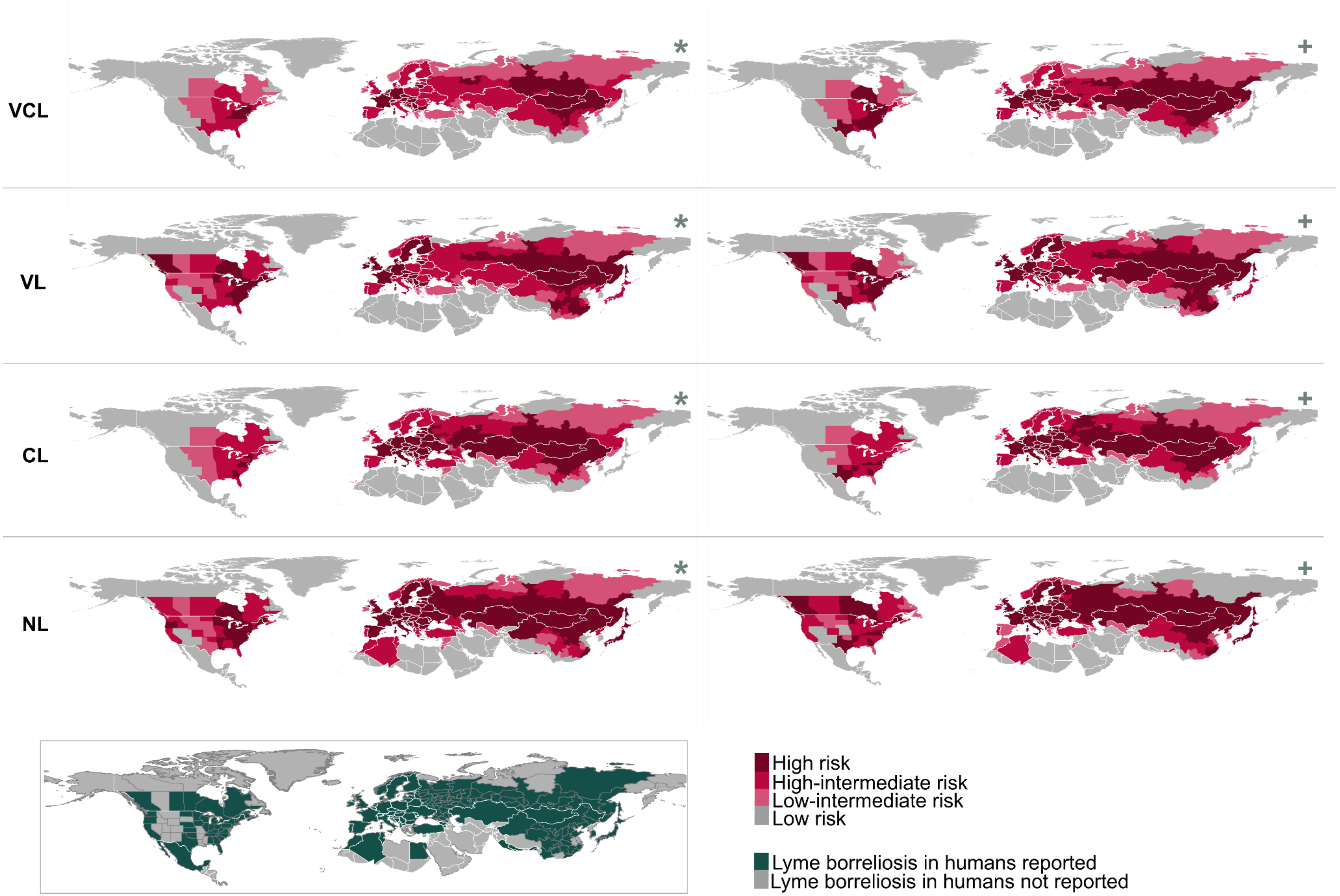
VCL
VL
CL
NL
*
+
High risk
High-intermediate risk
Low-intermediate risk
Low risk
Lyme borreliosis in humans reported
Lyme borreliosis in humans not reported

**Figure S4. Favourability models of tick species and their distribution within the study area.** Favourability values were categorized as low (<0.2), low-intermediate (0.2-0.5), high-intermediate (0.5-0.8), and high (>0.8).

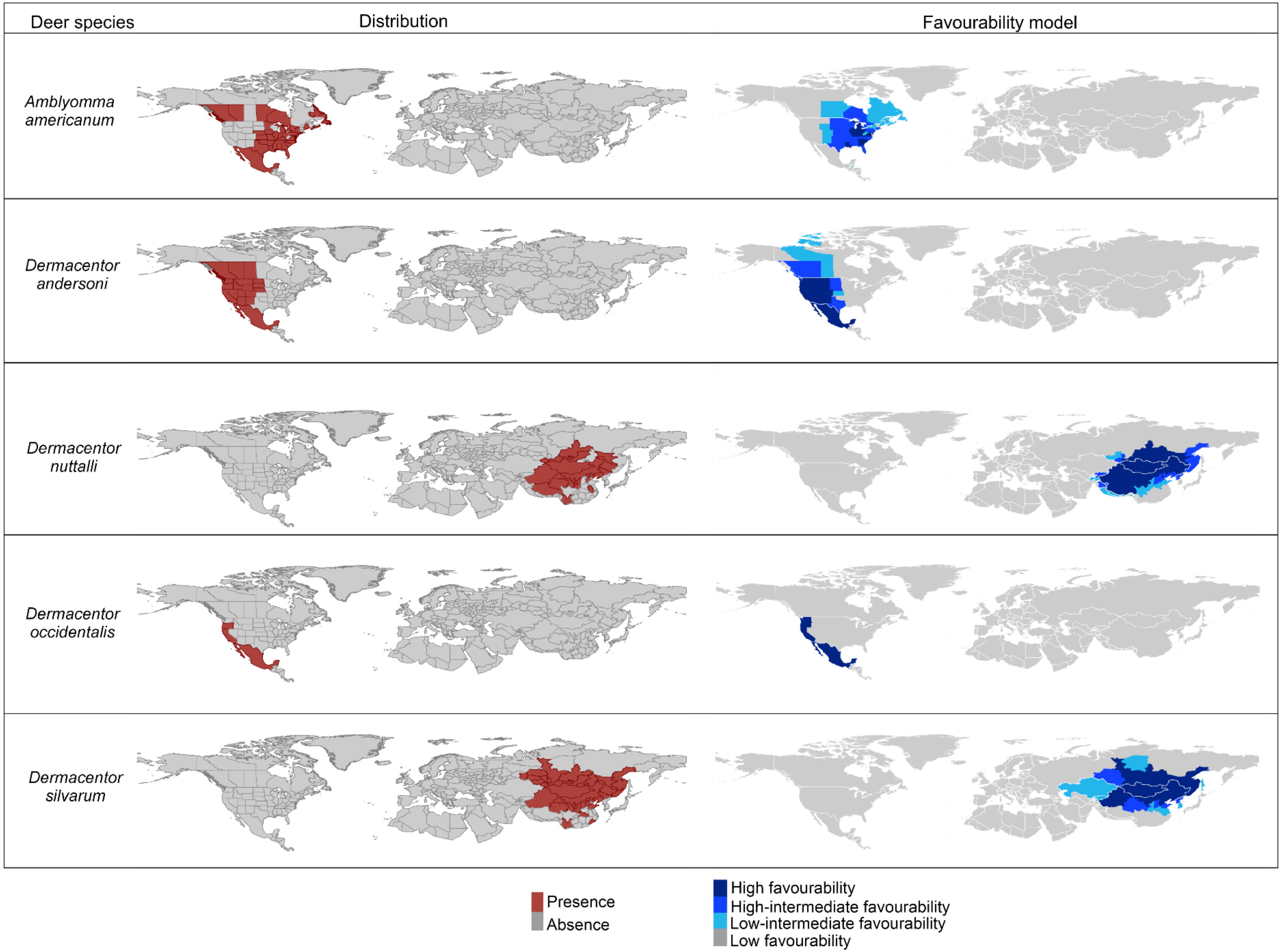

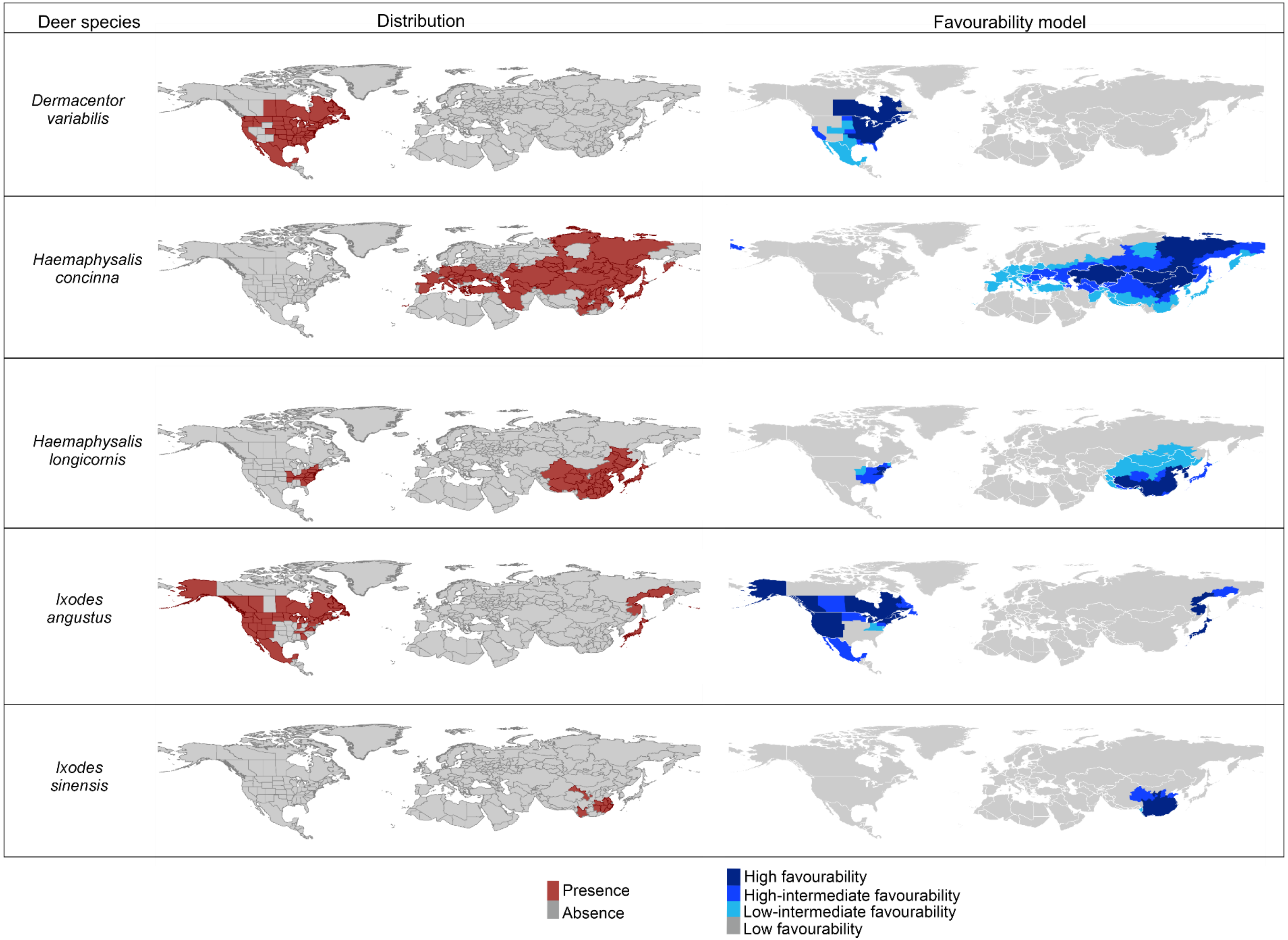
Deer species
Distribution
Favourability model
Dermacentor variabilis
Haemaphysalis concinna
Haemaphysalis longicornis
Ixodes angustus
Ixodes sinensis
Presence
Absence
High favourability
High-intermediate favourability
Low-intermediate favourability
Low favourability

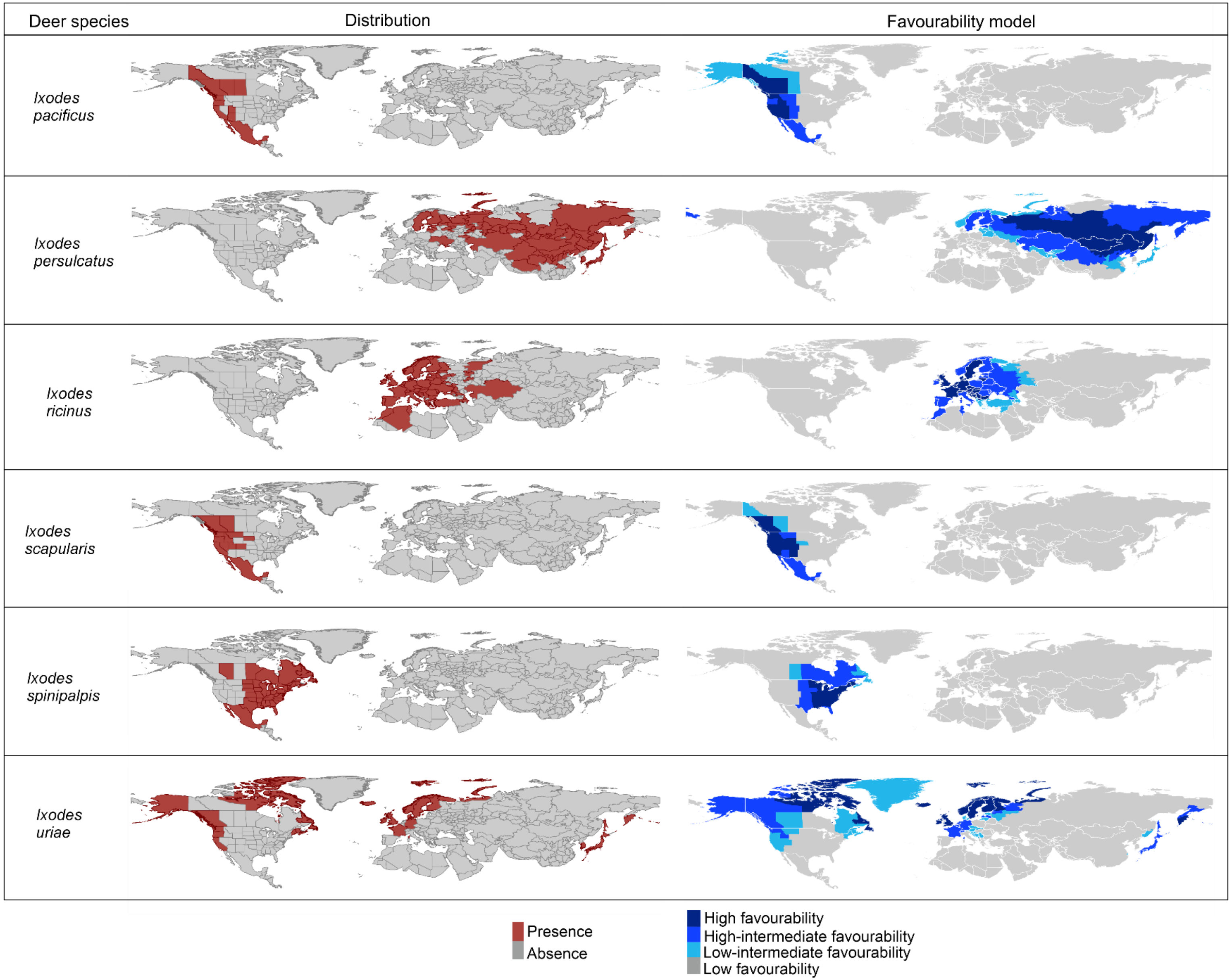
Deer species
Distribution
Favourability model
Ixodes pacificus
Ixodes persulcatus
Ixodes ricinus
Ixodes scapularis
Ixodes spinipalpis
Ixodes uriae
Presence
Absence
High favourability
High-intermediate favourability
Low-intermediate favourability
Low favourability

**Figure S5. Favourability models of deer species and their distribution within the study area.** Favourability values were categorized as low (<0.2), low-intermediate (0.2-0.5), high-intermediate (0.5-0.8), and high (>0.8).

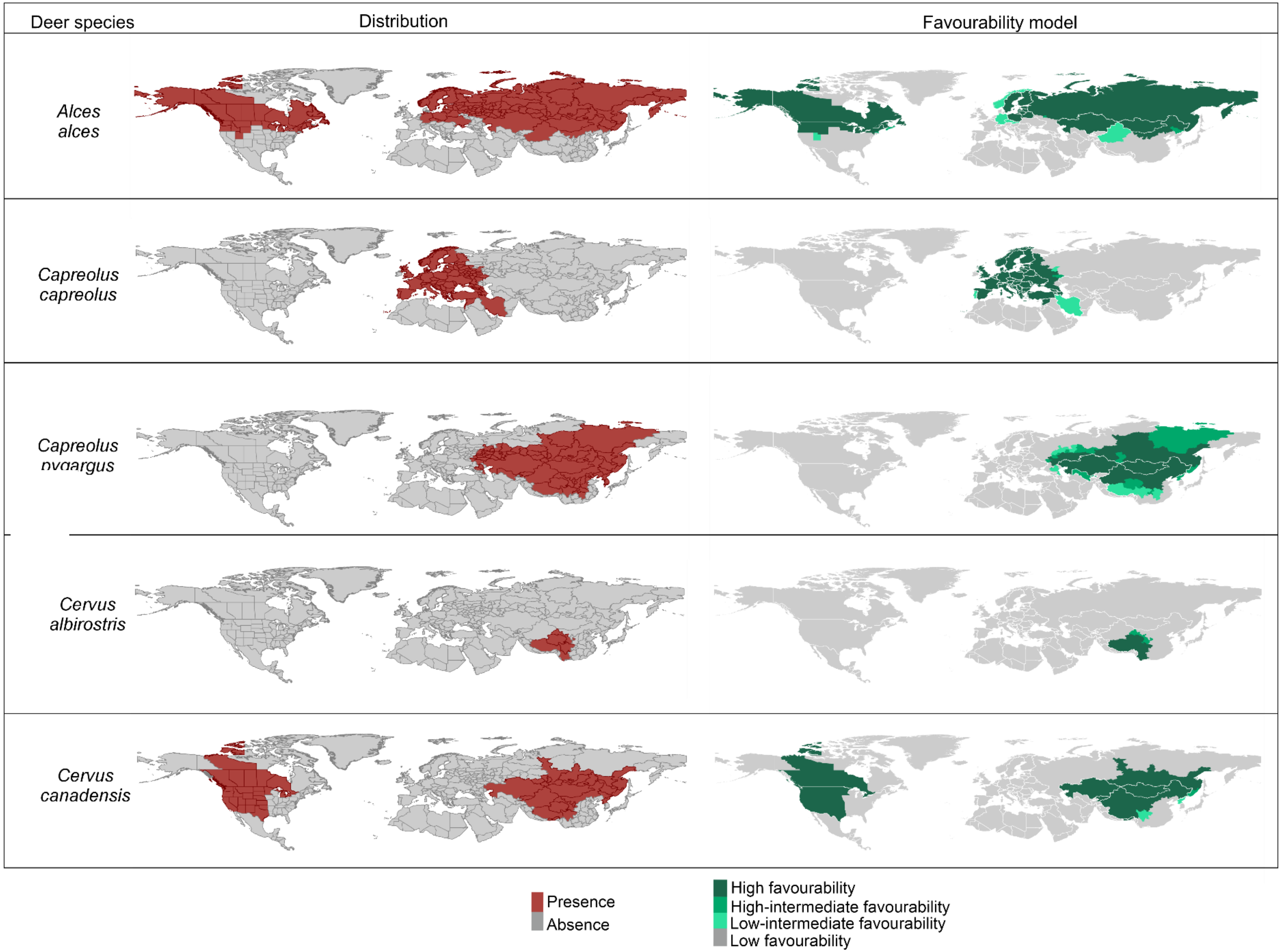

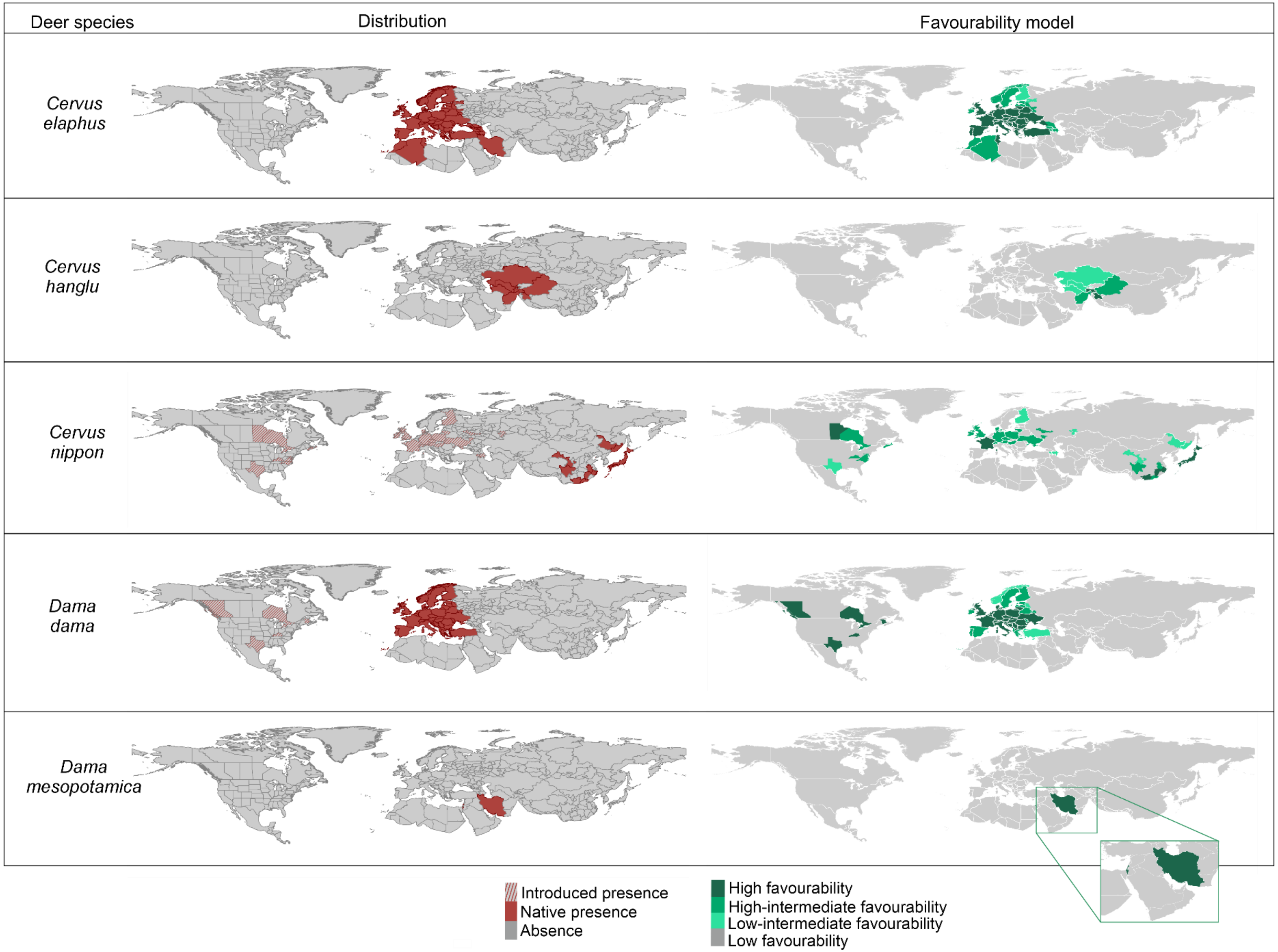

Deer species
Distribution
Favourability model
Cervus elaphus
Cervus hanglu
Cervus nippon
Dama dama
Dama mesopotamica
Introduced presence
Native presence
Absence
High favourability
High-intermediate favourability
Low-intermediate favourability
Low favourability

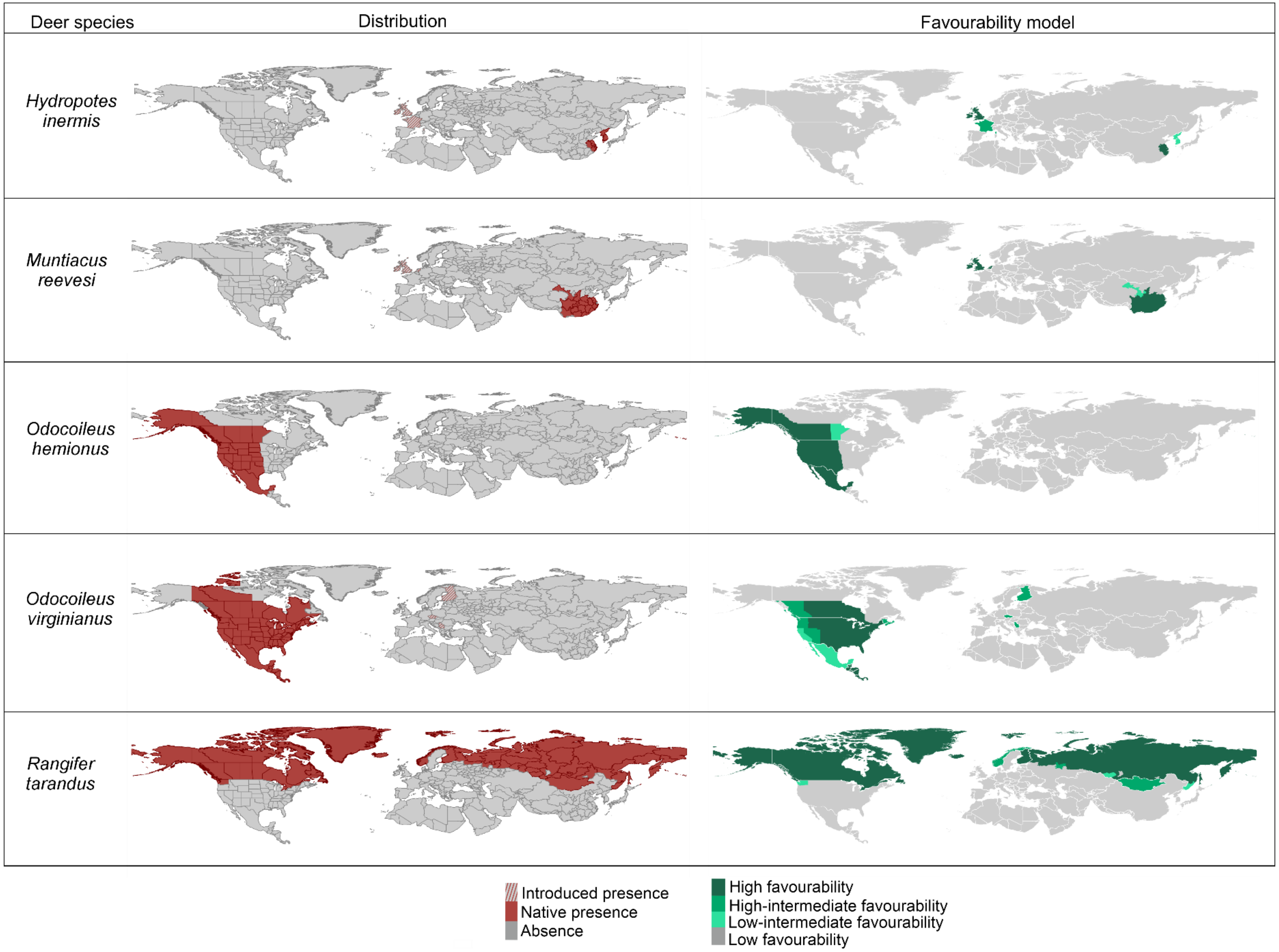
Deer species
Distribution
Favourability model
Hydropotes inermis
Muntiacus reevesi
Odocoileus hemionus
Odocoileus virginianus
Rangifer tarandus
Introduced presence
Native presence
Absence
High favourability
High-intermediate favourability
Low-intermediate favourability
Low favourability
